# Generating Synthetic Multi-national Longitudinal Cohorts for Clinically Grounded HIV Research

**DOI:** 10.1101/2025.11.14.25340245

**Authors:** Zhuohui J. Liang, Zhuohang Li, Nicholas J. Jackson, Yanink Caro-Vega, Ronaldo I. Moreira, Fabio Paredes, Jordany Bernadin, Diana Varela, Carina Cesar, Alessandro Blasimme, Jessica M. Perkins, Amir Asiaee, Stephany N. Duda, Bradley A. Malin, Bryan E. Shepherd, Chao Yan

**Affiliations:** Department of Biostatistics, Vanderbilt University Medical Center, Nashville, TN, USA; Department of Computer Science, Vanderbilt University, Nashville, TN, USA; Department of Biomedical Informatics, Vanderbilt University Medical Center, Nashville, TN, USA; Departamento de Infectología, Instituto Nacional de Ciencias Médicas y Nutrición Salvador Zubirán, Ciudad de México, México; Fundação Oswaldo Cruz (INI-Fiocruz), Instituto Nacional de Infectologia Evandro Chagas, Rio de Janeiro, Brazil; Facultad de Matemáticas, Pontificia Universidad Católica de Chile, Santiago, Chile; Département de l’Ouest, Les Centres GHESKIO, Port-au-Prince, Haiti; Instituto Hondureño de Seguridad Social, Tegucigalpa, Honduras; Fundación Huésped, Buenos Aires, Argentina; Department of Health Sciences and Technology, ETH Zurich, Zurich, Switzerland; Peabody College, Vanderbilt University, Nashville, TN, USA

## Abstract

High-quality, widely accessible international longitudinal cohort data for people living with HIV (PWH) have long been needed for advancing open science and data-driven innovation, yet stringent and incongruent privacy regulations have made data sharing difficult. Synthetic data generation offers a promising privacy-preserving alternative, but producing realistic synthetic cohorts of PWH remains challenging due to complex temporal dynamics, interdependent clinical variables, long follow-up periods, and high missingness inherent in such data. Here, we introduce Medical Longitudinal latent Diffusion (MeLD), a generative model designed to synthesize variable-length, decades-spanning, mixed-type clinical trajectories with missingness. Using the Caribbean, Central, and South America Network for HIV Epidemiology (CCASAnet) cohort, one of the world’s largest international HIV datasets with over 30 years of follow-up on nearly 50,000 PWH, we show that MeLD consistently outperforms state-of-the-art methods across data utility, fidelity, and privacy. Notably, MeLD excels in longitudinal inference utility, accurately reproducing time-to-death estimates and risk factor effects, while maintaining strong privacy protection. This work delivers the first in-depth, large-scale, and openly accessible synthetic longitudinal cohort of PWH that faithfully preserves the distributional patterns and clinical associations observed in real data, offering an immediately deployable resource for hypothesis generation, methods innovation, medical training, and reproducible HIV research.

## Introduction

Longitudinal clinical cohort data are invaluable for tracking the evolving needs and outcomes of individuals with chronic conditions. For people living with HIV (PWH), long-term clinical cohorts have been central to major advances in treatment development and epidemiological surveillance^1^. Continued progress in these areas hinges on the ability to share rich, individual-level clinical trajectories collected from diverse global sources. However, stringent and incongruent privacy regulations, ranging from the European Union’s General Data Protection Regulation (GDPR) to Brazil’s Lei Geral de Protecao de Dados (LGPD), have erected a patchwork of firewalls that hinder data sharing^2^. Even the most successful international research consortia approve only a handful of narrowly scoped data requests each year, slowing knowledge discovery and dissemination, and ultimately delaying clinical benefits.

Synthetic health data have emerged as a promising way to enable data sharing and collaboration across countries. When generated properly, synthetic data can mimic the statistical properties of source databases without linking produced records to real people, offering a pragmatic path to easier data sharing that preserves privacy^3–6^. Generative AI technologies have made it possible to produce realistic cross-sectional snapshots of electronic health record (EHR) data^7^ and simple longitudinal sequences^8^, enabling critical applications, such as trial-based association studies^9^, outcome predictive modeling^10,11^, geotemporal epidemic forecasting^12^, and DNA-disease association analysis^13^—while researchers navigate complex administrative processes required to access real data.

However, generating high-quality synthetic longitudinal clinical cohort data faces non-trivial challenges. Cohorts of people with chronic conditions like HIV typically span decades and include critical time-to-event outcomes, such as a variety of clinical endpoints (e.g., diagnoses of comorbidities), medication changes, treatment failures, and mortality. These complexities are fundamental to ensuring clinical relevance and usability of longitudinal data. Yet current synthetic data generation methods struggle to capture the temporal depth and clinical complexity of chronic condition trajectories. Most of these methods are developed and evaluated with short horizons (e.g., 24-hour ICU streams or datasets with a limited number of visits^8^), which can reduce a marathon-length chronic condition journey to a snapshot. Another challenge arises from the need to process and generate a mix of continuous (e.g., HIV viral load measurements), discrete (e.g., test frequencies), and categorical (e.g., diagnostic codes) variables, while simultaneously preserving their temporal relationships. Existing methods, mostly adapted from unimodal domains like imaging or natural language processing, rely on tokenizing continuous and discrete values into coarse categories. This process potentially obscures clinically meaningful variations^14^. Additionally, current synthetic data evaluation strategies typically rely on generic resemblance metrics^15^. These are limited in scope. Specially, they do not assess whether scientifically relevant relationships are preserved in synthetic cohort data, which would be a far more demanding standard of data quality. For instance, an essential but missing metric is whether models fitted on synthetic data can reproduce time-to-event patterns observed in real cohorts, an outcome that reflects the analytical value of longitudinal clinical cohort data and is essential for evaluating treatment effects and informing health policy. Another overlooked metric examines whether longitudinal synthetic data can yield risk factor estimates comparable to those derived from real data, which addresses the capability of synthetic data to support scientific hypothesis generation.

To address these challenges, we introduce **Me**dical **L**ongitudinal latent **D**iffusion (MeLD), a longitudinal latent diffusion model built on a transformer backbone (Fig. 1). At a high level, MeLD first embeds variables of mixed types into a continuous latent space via a variational autoencoder (VAE), which naturally preserves numerical precision and handles inter-visit time gaps. A transformer-powered diffusion process then learns dependencies across timescales, ranging from short-term oscillations in laboratory measurements to multi-decade survival trends within the latent representations. We complement MeLD with evaluation metrics that jointly assess data utility, fidelity, and privacy, with particular emphasis on time-to-event analysis integrity and risk factor inference reproducibility.

**Figure 1.**
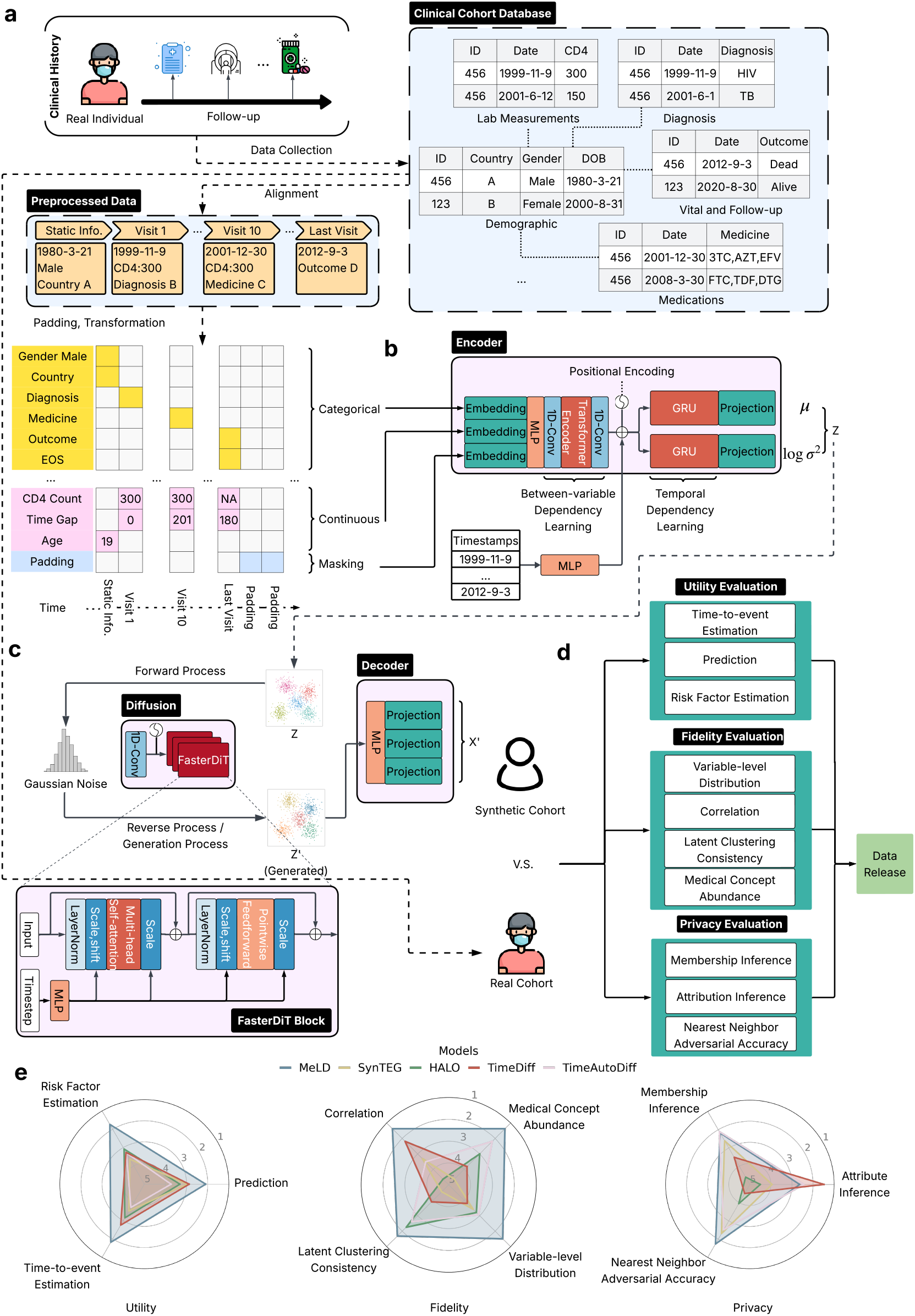
An overview of the MeLD architecture, data quality evaluation framework, and corresponding overall model rankings. **a**, Individual-level information from demographics, vital and follow-up status, visits, laboratory results, medications, and diagnoses is time-aligned and structured into a visit-centric sequence, then padded to create AI-ready data. **b**, Visit-level clinical trajectories, containing mixed continuous and categorical fields, are embedded, projected, then passed to a VAE encoder to produce their latent representations. **c**, A DiT-based diffusion module generates synthetic clinical trajectories in the latent space by sampling from Gaussian noise, which are then decoded by the VAE decoder back into the original data space, yielding realistic longitudinal records of variable length. **d**, The resulting synthetic data undergo systematic evaluation across data utility, fidelity, and privacy assessment prior to release. **e**, Ranks of MeLD across data utility, fidelity, and privacy, compared to state-of-the-art synthetic longitudinal data generation methods. Model rankings in each dimension are derived across ten synthetic datasets, generated by two independently trained models, with five datasets generated from each. Smaller values in ranks correspond to higher performance. VAE=variational auto-encoder; MLP=multi-layer perceptron; DiT=diffusion transformer; GRU=gated recurrent units.

We demonstrate MeLD by generating the first large-scale, shareable, longitudinal synthetic cohort of PWH based on the Caribbean, Central, and South America network for HIV Epidemiology (CCASAnet) cohort^16,17^, which is part of the International epidemiological Databases to Evaluate AIDS (IeDEA) network—the world’s largest HIV research consortia. The CCASAnet data used in this study pool longitudinal medical records and routinely collected data to track approximately 50,000 PWH across six countries, with follow-up spanning over 30 years. Systematic assessment demonstrates that MeLD produces synthetic cohort of PWH that closely mirror the real records across data utility and fidelity metrics, outperforming state-of-the-art generative methods, while remaining robust to standard privacy inference. Notably, MeLD achieves superior performance in preserving long-term time-to-event patterns and risk factors for mortality and clinical endpoints seen in real records. The shareable synthetic replica of the CCASAnet cohort represents a significant step forward in enabling HIV data sharing and opens new avenues to generate meaningful insights that can drive HIV-focused training, education, research, and real-world applications.

## Results

### An overview of MeLD and experimental setup

#### Longitudinal clinical cohort data

This study relies on CCASAnet, a U.S. National Institutes of Health (NIH)-funded multi-country consortium that harmonizes routine clinical care data from HIV clinics in Argentina, Brazil, Chile, Haiti, Honduras, Mexico, and Peru. As of December 2024, the cohort (excluding those from Peru) comprises 49,606 adults ( ≥18 years) living with HIV. The CCASAnet database, like those of many chronic condition cohorts, follows a normalized, relational design. Immutable variables (e.g., demographics) sit in cross-sectional tables, whereas time-varying data (e.g., timestamped visits, laboratory results including CD4 cell count and HIV viral load measurement, medications including antiretroviral therapy regimens with start and stop dates, and clinical diagnoses) are in longitudinal tables. This structure is common because it mitigates redundancy, allows incremental updates, and enables analysts to assemble cohorts by joining relevant tables. However, raw data require systematic preprocessing to be suitable for model fitting. To enable longitudinal modeling for synthetic data generation, we reorganize the CCASAnet data into a visit-centric format, where longitudinal data are grouped by clinical visits. To balance comprehensive coverage with computational feasibility, we retain up to 120 visits per person, corresponding to the 90th percentile and approximately 36 years of maximum follow-up. Inter-visit time gaps are derived and added as a continuous variable. The resulting real analytic dataset averages 45 visits per person and includes 135 variables. Full details of the datasets are provided in the Methods Section.

#### Architectural design of MeLD

After assembling longitudinal records into visit-level sequences prefixed by demographic information, an end-of-sequence (EOS) token is appended to each person’s final visit. Right-padding is applied with zero-filled pseudo-visits up to a fixed horizon (Fig. 1a). The resulting data preserve chronological order and data type heterogeneity while matching the fixed-size input expected by the encoder of MeLD. Two binary mask sequences with the same length as the constructed data are then created: one flags padded pseudo-visits, the other flags missing values. See the Methods Section for data preprocessing details. MeLD adopts a latent diffusion architecture, in which a VAE translates visit-level representations between the original and latent spaces (Fig. 1b,c). Within the latent space, a diffusion model generates synthetic visit sequences, avoiding the complexity of handling diverse data types in the original space (Fig. 1c). Specifically, the VAE encoder of MeLD first embeds categorical values, continuous values, and data masks (from missing data and padded pseudo-visits), into their own representations using stream-specific embedding modules. At each visit, these embeddings are concatenated and fused through a multi-layer perceptron (MLP) to form a unified representation. To capture the fine-grained associations across variables, we apply multi-channel 1D-convolutions before and after a transformer encoder that models variable-wise dependencies^18^. Temporal dynamics across visits are then modeled and aligned using two additional gated recurrent units (GRU), conditioned on embedded timestamps. The output of the VAE encoder is a sequence of temporally aligned visit-level latent representations that serves as the foundation for the subsequent diffusion process, enabling realistic and coherent synthesis of longitudinal records. These representations are parameterized by the mean (*μ*) and variance (*σ* ^2^) of the posterior distribution of each person’s underlying clinical trajectory in the latent space.

MeLD adopts diffusion transformer (DiT)^19^ as its generative backbone, a state-of-the-art algorithm developed for synthetic image generation that replaces the traditional U-Net architecture with a stack of transformer blocks. For longitudinal clinical visit sequences, we adapt DiT by representing each latent visit as a token and performing denoising entirely in the latent space. Gaussian noise is progressively removed from lower-quality visit sequences using self-attention blocks, allowing the model to generate entire clinical trajectories simultaneously at each denoising step, rather than performing autoregressive generation. To address the notoriously long training cycles of standard diffusion, we integrate the FasterDiT acceleration scheme^20^, which refines training supervision by guiding the model to match both the size and the direction of each denoising velocity step to improve convergence speed. In the generation stage, synthetic clinical visit sequences are produced in the latent space by iteratively denoising Gaussian noise into realistic trajectories.

MeLD reconstructs synthetic longitudinal records by decoding the denoised latent sequences using the VAE decoder composed of stacked MLP layers. Finally, three independent projection modules map the decoded sequences into categorical, continuous, and mask fields (missing and EOS masks). We then convert these sequences to the original data format.

#### Experimental setup

We conduct evaluations at two levels. First, we benchmark MeLD against representative state-of-the-art longitudinal clinical data generators, including models based on generative adversarial network (GANs), language models, diffusion, and latent diffusion. Second, we compare standard MeLD with its variants that swap in alternative diffusion and VAE architectures to assess the impact of MeLD design choices on overall data quality. Specifically, we include the following models:

1. SynTEG^21^: This model generates timestamped diagnosis sequences through a two-stage process that decouples temporal modeling from visit generation. In the first stage, a recurrent neural network models visit-level dynamics and temporal dependencies in real data by jointly predicting the diagnoses at the next visit and the inter-visit time interval. In the second stage, the hidden states learned from the temporal model serve as the condition of a Wasserstein GAN, which samples the multivariate distribution of diagnoses for each visit. SynTEG does not natively support mixed-type data, so we discretize continuous variables into quantile-based bins, convert them into categorical representations for generation, and generate synthetic continuous values through uniform sampling within relevant bins.
2. HALO^22^: This is a hierarchical language model that decomposes dependency learning into two levels: a coarse-grained, visit-level module and a fine-grained, code-level module. HALO generates visit representations and variable values within each visit autoregressively until an EOS token is produced. HALO also lacks native support for mixed-type data; continuous variables are discretized for training and later generated through sampling using the approach described above for SynTEG.
3. TimeDiff^8^: This model separately performs Gaussian diffusion for continuous variables and multinomial diffusion for categorical variables, both in the original space. A time-conditioned bi-directional long short-term memory (LSTM) network parameterizes the diffusion process, capturing sequence dynamics in one pass, rather than autoregressively. Since TimeDiff is not designed to generate variable-length sequences, we determine the endpoint of each generate sequence based on the occurrence of death or censoring.
4. TimeAutoDiff^23^: This model uses a latent diffusion architecture with a two-stage training process similar to MeLD. In the first stage, a VAE is trained to project full-length clinical visit sequences between the original mixed-type data space and the latent space. In the second stage, an LSTM-based denoising diffusion probabilistic model (DDPM)^24^ is trained to reconstruct latent trajectories from Gaussian noise. Noticeably, real timestamps are used both as conditions for second-stage model training and as the “synthetic” timestamps, which increases vulnerability to privacy intrusions and constrains data diversity. To enable a head-to-head comparison with MeLD, we instruct TimeAutoDiff to generate synthetic inter-visit intervals. TimeAutoDiff also does not have an mechanism for variable-length generations, so like TimeDiff, we use death and censoring as terminal events for each generated sequence.

To assess the impact of MeLD’s diffusion module on synthetic data quality, we substitute it with the following alternative diffusion architectures for modeling temporal dependencies, while keeping all other components unchanged: 1) DDPM, a Gaussian diffusion model with a convolutional U-Net denoiser^24^, 2) LSTM, the recurrent backbone used in TimeAutoDiff, modified by removing real timestamp conditioning to be consistent with MeLD’s design, and 3) DiT, a transformer-based denoiser without the convergence acceleration strategies employed in MeLD^19^. We also evaluate the impact of MeLD’s VAE component by comparing it to a model that substitutes both MeLD encoder’s GRU and decoder’s MLP with a transformer. This is to test whether a stronger temporal model can bring significant benefit to synthetic data quality. We refer to these baseline models as MeLD-DDPM, MeLD-LSTM, MeLD-DiT, and MeLD-Transformer, respectively. As MeLD-DiT needs substantially longer time to converge, we constrain its training time to match that of MeLD.

We randomly partition the CCASAnet cohort into training (85%) and test sets (15%). Each model is trained twice independently on the training set (i.e., two runs), with five synthetic cohorts generated for each run. This yields ten synthetic datasets, all matching the size of the real training set. These ten datasets are used to estimate the 95% confidence intervals (CI) for evaluation metrics using a normal approximation. Since MeLD is the only model explicitly designed to handle missing data in model training and generation, we impute missing values in the real data beforehand using Multivariate Imputation by Chained Equations (MICE)^25^, a standard approach in epidemiology research, and perform evaluation against imputed data. For MeLD, missing values in the generated synthetic data are post-processed using the same imputation pipeline applied to the real data for consistency in evaluation.

We evaluate the quality of the synthetic data along three complementary dimensions (Fig. 1d): 1) data utility, 2) data fidelity, and 3) privacy risks. Recognizing that existing evaluations overemphasize generic resemblance scores and simple outcome prediction while overlooking whether scientifically meaningful relationships are preserved, we explicitly test if models fitted on synthetic data can recover key longitudinal phenomena that are central to HIV research. In the utility dimension, we investigate the utility of the synthetic data to develop prediction models for clinical endpoint prediction. We then go beyond this standard evaluation and investigate time-to-event analysis and risk factor estimation, considering whether time-to-event relationships and covariate effects observed in the real cohort are reproducible in synthetic data. Data fidelity analyses investigate whether statistical properties of the data are preserved, at the variable-level, individual-level, and across time. This is quantified through variable-level distribution and correlation resemblance, latent clustering consistency, medical concept abundance, and missingness patterns. Finally, we apply standard privacy metrics to gauge the risks in sharing the synthetic data.

### Data Utility

To evaluate data utility, we select key tasks that reflect priorities of the HIV research community and the core value of longitudinal clinical cohorts, including time-to-event estimation, outcome prediction, and risk factor estimation.

#### Time-to-event estimation

A generative model must capture both short- and long-term dependencies in real data to produce realistic time-to-event estimates. Unlike common evaluation metrics that focus on individual variables or pairwise relationships, time-to-event estimation relies on derived variables computed from multiple underlying fields, making it more sensitive to compounding errors that can occur in synthetic data. Therefore, accurately reproducing time-to-event curves presents a significantly greater challenge for generative models than matching simple statistics alone. We estimate cumulative hazard functions using the Nelson-Aalen method and quantify Kaplan-Meier distance (KM-D) between synthetic and real data^26^. Two-sided log-rank tests compare survival functions derived from the real and synthetic data. Synthetic and real training data of the same sample size are compared against real test data to allow comparability of *p*-values. Additionally, we evaluate the maximum follow-up time derived from synthetic datasets.

In HIV epidemiology, time from antiretroviral therapy (ART) initiation to death is a central outcome, reflecting treatment efficacy, retention in care, and mortality risk. Fig. 2a-c presents aggregated results across all ten synthetic datasets (see detailed results in Supplementary Table 3), while Fig. 2d-l shows the Nelson-Aalen cumulative hazard functions (i.e., survival curves) estimated from real training and test data, as well as from one representative synthetic dataset generated by each model, selected to be closest to the mean of KM-D results for illustrative purposes. There are multiple notable findings here. First, the cumulative hazard functions estimated from MeLD-generated synthetic data exhibit the closest agreement with that derived from real data (Fig. 2a,b), yielding the largest *p*-value (mean [95% confidence interval]: 0.815 [0.521, 1.000]) and the smallest KM-D (0.015 [0.005, 0.024]), as illustrated by the example in Fig. 2d. The other models produce remarkably smaller *p*-values and larger KM-D. Second, current models display systematic biases by either 1) overestimating early death (e.g., HALO, Fig. 2f) or 2) underestimating later death to varying degrees (e.g., SynTEG and TimeDiff, Fig. 2e,g). TimeAutoDiff, in contrast, exhibits the greatest variability in its time-to-death estimates, indicating reduced stability (Fig. 2a,b). Third, all of the models generate maximum follow-up time ranges longer than that observed in real data (Fig. 2c). However, MeLD produces the range closest to that of real data. By contrast, SynTEG and TimeDiff yield follow-up ranges more than double the actual range (36 years). Fourth, MeLD variants with alternative diffusion modules (MeLD-DDPM, MeLD-LSTM, and MeLD-DiT) all demonstrate inferior performance than the standard MeLD, whereas MeLD-Transformer, despite its enhanced capability for modeling temporal dependencies in VAE, does not lead to better time-to-death estimates. This highlights the advantages of the MeLD architecture in faithfully reproducing time-to-death estimation.

**Figure 2.**
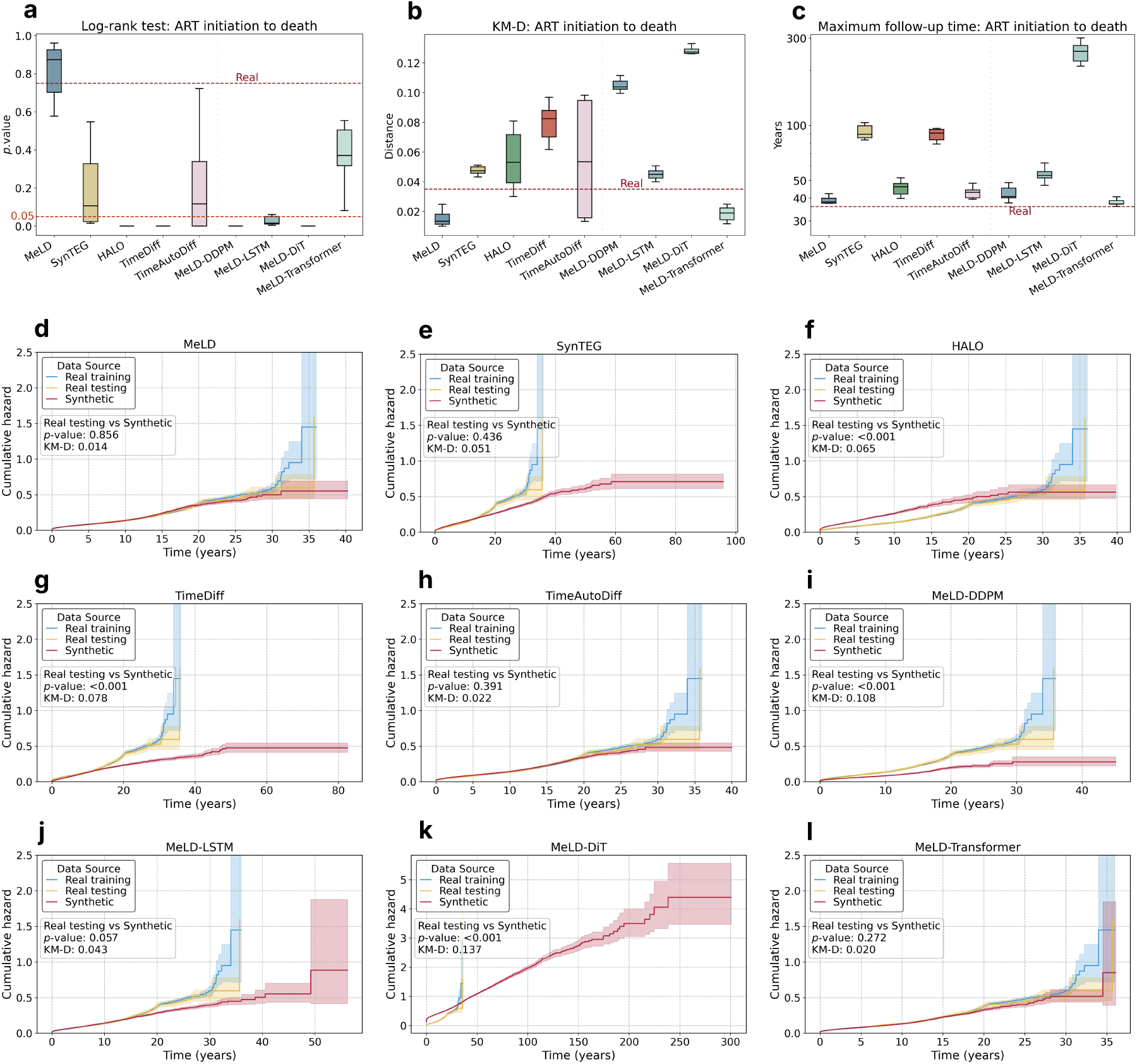
Comparison of performance in time-to-death estimation. Time from ART initiation to death is evaluated using: **a**, log-rank test *p*-values, **b**, Kaplan-Meier distance (KM-D), and **c**, maximum follow-up time, with real test data as the reference. Results based on real training data serve as the baseline, shown with red dashed lines. Boxplots are made with ten synthetically generated datasets. **d**-**l**, estimated Nelson-Aalen cumulative hazard functions of real training and test data, along with one synthetic dataset generated by each model, selected to be closest to the mean of KM-D values. KM-D between survival functions estimated from real test and synthetic data, as well as *p*-values from two-sided log-rank tests comparing estimated survival functions of real test and synthetic data are indicated.

We also evaluate the utility of synthetic data in reproducing the time-to-event analyses from ART initiation to the first diagnosis event across 50 regularly collected clinical endpoints, encompassing conditions such as tuberculosis, diabetes, and bone diseases. We conduct the same evaluation as in the time-to-death analysis, adding a Bonferroni correction to the log-rank tests to account for multiple comparisons. We summarize results across the 50 clinical endpoints using 1) the false discovery rate (FDR), calculated as the ratio of clinical endpoints where synthetic data exhibit significant differences, and 2) the distribution of KM-D values.

MeLD and TimeDiff achieve the lowest FDR and KM-D values (Table 1). Moreover, all MeLD variants perform worse than the standard MeLD in terms of FDR and KM-D values, with the exception of MeLD-Transformer, which yields comparable KM-D values. These results support the same conclusion as the time-to-death estimations regarding the advantage of the MeLD architecture. Further, the clinical endpoints most frequently misrepresented in time-to-diagnosis estimates vary in their prevalence across models. MeLD and TimeDiff tend to misrepresent clinical endpoints with lower prevalence (<10%), whereas other models are more likely to misrepresent those with higher prevalence (>10%), which are generally easier to model. Taken together the time-to-death and time-to-diagnosis results, MeLD demonstrates the greatest data utility for time-to-event estimation among all the models, whereas TimeDiff struggles to model longer-term dependencies like death.

**Table 1.** Performance comparison in time-to-diagnosis estimation across 50 clinical endpoints. False discovery rate (FDR) from log-rank tests and KM-D values are derived using real test data as the reference. Results based on real training data serve as the baseline. 95% confidence intervals are reported over ten synthetically generated datasets. Best-performing results are highlighted in bold for comparisons between MeLD and existing models, as well as among MeLD variants. Top three clinical endpoints that are worst estimated are provided in descending order of their KM-D values, as well as their prevalence in real data.

| Model | FDR from log-rank tests | KM-D distribution | Top-3 errors | Prevalence |
| --- | --- | --- | --- | --- |
| Real training (vs. real test) | 0 | 0.001 [0.000, 0.004] | - | - |
| MeLD | 0.190 [0.144, 0.236] | **0.003 [0.000, 0.014]** | Diabetes<br>Bone disease<br>Dengue | 2.09%<br>6.09%<br>0.83% |
| SynTEG | 0.312 [0.259, 0.365] | 0.006 [0.000, 0.035] | Dyslipidemia<br>Diabetes<br>Dengue | 10.60%<br>2.09%<br>0.83% |
| HALO | 0.212 [0.156, 0.268] | 0.010 [0.000, 0.075] | Dyslipidemia<br>Tuberculosis<br>Cancer | 10.60%<br>15.47%<br>3.34% |
| TimeDiff | **0.100 [0.068, 0.132]** | **0.003 [0.000, 0.013]** | Candidiasis pulm<br>Pneumocystis<br>Cytomegalovirus | 1.12%<br>3.67%<br>0.80% |
| TimeAutoDiff | 0.606 [0.536, 0.632] | 0.007 [0.000, 0.036] | Dyslipidemia<br>Tuberculosis<br>Diabetes | 10.60%<br>15.47%<br>2.09% |
| MeLD-DDPM | 0.554 [0.487, 0.621] | 0.011 [0.000, 0.080] | Dyslipidemia<br>Tuberculosis<br>Diabetes | 10.60%<br>15.47%<br>2.09% |
| MeLD-LSTM | 0.300 [0.248, 0.352] | 0.006 [0.000, 0.050] | Tuberculosis<br>Dyslipidemia<br>Diabetes | 15.47%<br>10.60%<br>2.09% |
| MeLD-DiT | 0.936 [0.919, 0.953] | 0.079 [0.000, 0.266] | Tuberculosis<br>Cancer<br>Toxoplasmosis | 15.47%<br>3.34%<br>15.47% |
| MeLD-Transformer | **0.290 [0.225, 0.355]** | **0.003 [0.000, 0.016]** | Diabetes<br>Dyslipidemia<br>Dengue | 2.09%<br>10.60%<br>0.83% |

#### Prediction

To examine the prediction utility of MeLD-generated synthetic data, we choose three disease prediction tasks relevant to understanding disease progression for PWH, each with different prevalence and pathogenesis: the development of 1) tuberculosis, 2) cancer, and 3) cardiovascular complications after six months of program enrollment. We use data observed in the first six months as predictors to fit a simple one-layer GRU model. Predictors include demographics, weight, height, CD4 cell count, HIV viral load, ART regimens, and clinical diagnoses. PWH diagnosed before six months are excluded from the respective tasks. We evaluate predictive performance using AUROC on real test data, comparing training a classifier on synthetic data (TSTR) to training it on real training data (TRTR).

MeLD ranks among the top-performing models across all three prediction tasks (Fig. 3, Supplementary Table 4), with mean AUROC of 0.757 [0.741, 0.773], 0.626 [0.597, 0.655], and 0.615 [0.541, 0.689] for predicting tuberculosis, cancer, and cardiovascular disease, respectively. Notably, for cancer prediction, models trained on MeLD-generated synthetic data surpass those based on real training data. This might be attributed to the VAE component in MeLD, which removes noisy relationships in cancer development when learning latent representations, thereby generating synthetic data with reduced noise. Other models, by contrast, do not achieve consistently high rankings across tasks. For example, while TimeDiff exhibits top and moderate performance for cardiovascular disease and tuberculosis prediction, respectively, it is near the bottom for cancer prediction. HALO achieves the second best performance for cancer prediction, yet cancer is among the clinical endpoints that are most misrepresented in its time-to-event analysis (Table 1). This discrepancy suggests that time-to-event analyses and prediction tasks focus on complementary aspects of data utility and that reliable generative models should score well on both.

Among the MeLD variants, those using alternative diffusion modules exhibit lower prediction performance than the standard MeLD across all tasks. In contrast, MeLD-Transformer performs well for all prediction tasks, achieving an AUROC comparable to the TRTR scenario. Although MeLD-Transformer has a higher mean AUROC than MeLD, differences are minor, again suggesting that substituting the temporal component in VAE with a transformer provides only marginal advantages.

**Figure 3.**
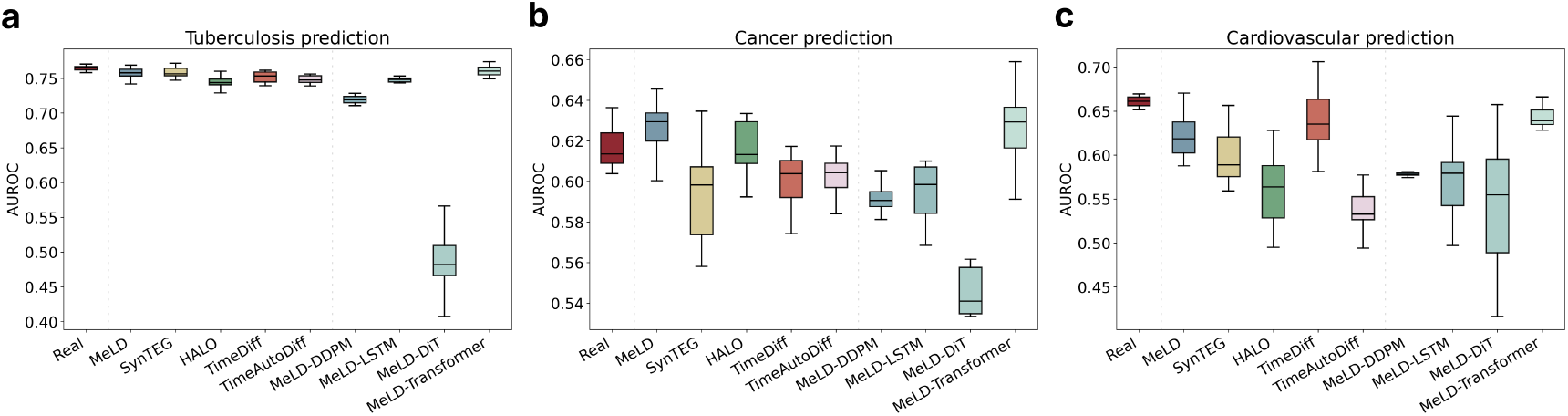
Comparison of performance in outcome prediction. Prediction performance on the real test data characterized by AUROC, is shown for **a**, tuberculosis, **b**, cancer, and **c**, cardiovascular diseases. Models are trained on data accumulated during the first six months post-enrollment to predict subsequent clinical endpoints. The scenario of training on real training data and testing on real test data serves as the baseline. Boxplots for synthetic data generation models are made with ten synthetically generated datasets, whereas those for training on real training data (TRTR) are based on ten random splits of the training and validation datasets.

#### Risk factor estimation

HIV researchers often estimate statistical associations between risk factors and clinical events to better understand disease etiology, identify high-risk subgroups, and develop prevention and treatment strategies. Thus, a critical test of synthetic data utility is whether it can preserve clinically meaningful associations. To examine this, we fit Cox proportional hazards models to synthetic data to estimate the association between risk factors at ART initiation and mortality^27^. We compare estimates to those derived from real data to evaluate both the quality of the synthetic data and its potential applicability for clinical and public health research. We include sex, age, CD4 cell count, and calendar year at ART initiation as covariates, with models stratified by study site^28^. For evaluation, we quantify three types of errors: 1) direction error: a significant estimate of a risk factor yields a log-hazard ratio with an opposite sign in the synthetic data compared to the significant estimate from real data; 2) type I error: a risk factor is significant in the synthetic data but not in the real data, and 3) type II error: a risk factor is significant in the real data but not in the synthetic data. We then compare the log-hazard ratio values and corresponding confidence intervals of risk factors estimated from real data and from MeLD-generated synthetic data.

Across all ten synthetic datasets and risk factors, MeLD produces the fewest errors for each type, as well as the lowest total error (Fig. 4a). Specifically, MeLD makes only one type I error, which occurs for the age risk effect (Fig. 4b), and incurs no type II or direction errors. This corresponds to an overall error rate of 2.5%. By contrast, all other models make errors across all three types. For example, HALO-generated synthetic data produce 15 total errors (the second fewest), yet substantially higher than MeLD. Other models exceed 37.5% total error rates, and all MeLD variants produce error rates *>*25.0%. While MeLD produces the fewest errors, some of its estimates are biased compared to those from real data (Fig. 4b). For example, although MeLD introduces no direction, type I, or type II errors for the calendar year log-hazard ratio, there is no confidence interval overlap between any of MeLD’s ten synthetic datasets and that from the real data. More details can be found in Supplementary Table 5.

**Figure 4.**
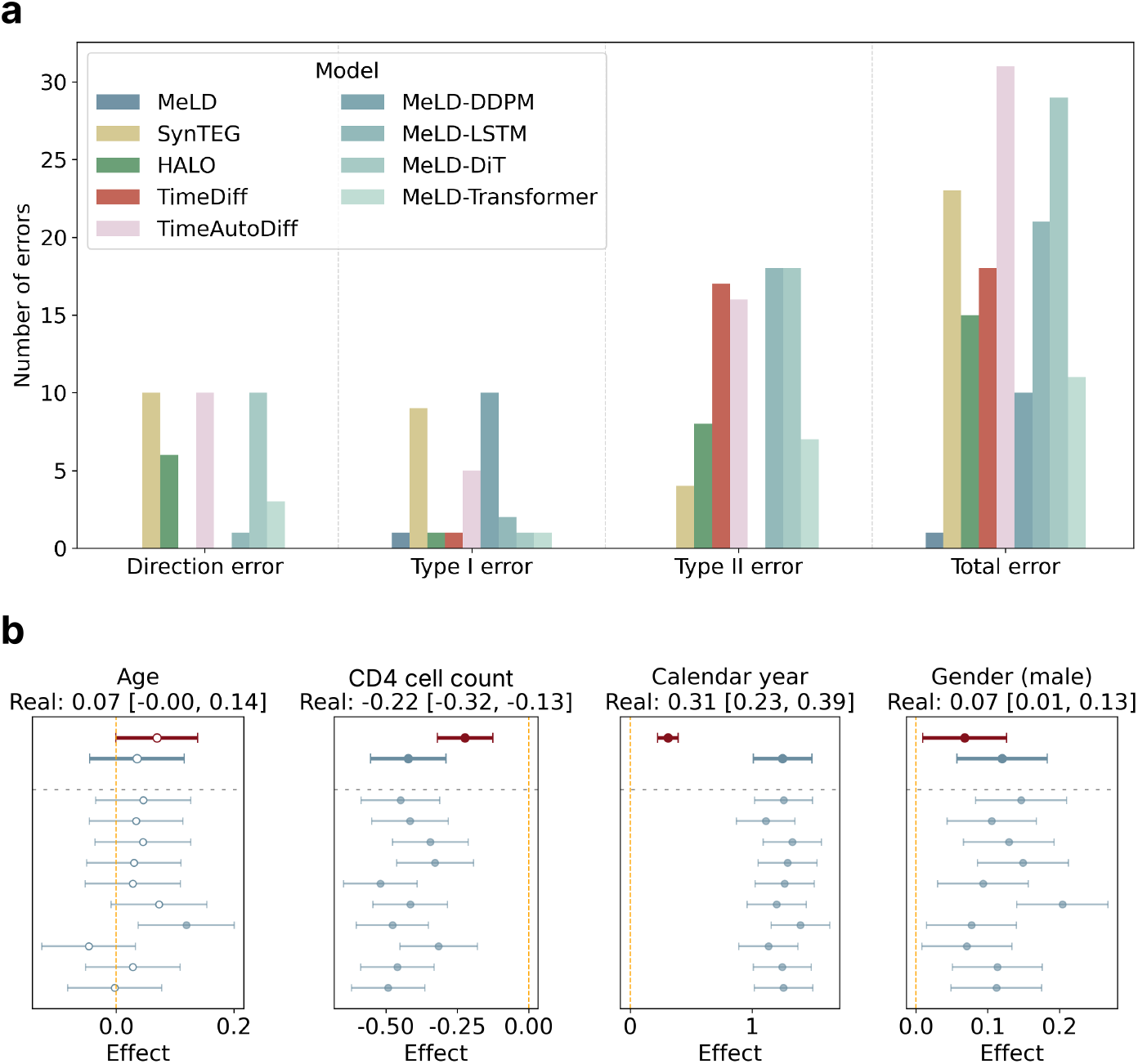
Error analysis for risk factor estimation. **a**, Aggregated errors from Cox proportional hazard models evaluated on ten synthetic datasets across four risk factors. A direction error occurs when a significant estimate of a risk factor yields a log-hazard ratio with an opposite sign in the synthetic data compared to the real data; a type I error occurs when a risk factor is identified as significant in the synthetic data but not in the real data; a type II error occurs when a risk factor that is significant in the real data is missed in the synthetic data. **b**, Forest plots of log-hazard ratio risk factor estimates from MeLD’s ten synthetic datasets and their aggregated estimates using Rubin’s Rule (bold blue), alongside estimates from real data (bold red). Non-significant estimates are indicated using hollow-centered circles.

### Data Fidelity

Data fidelity focuses on evaluating the statistical properties of synthetic data that are independent of specific applications and determining how well they mirror those of real data. High-fidelity synthetic longitudinal cohort data should demonstrate close resemblance in terms of variable-level distributions, correlations, latent clustering consistency, individual-level medical concept abundance, and overall missingness patterns.

#### Variable-level distribution

For continuous variables, we assess: 1) total number of visits, 2) inter-visit time gaps, 3) weight, 4) height, 5) enrollment age, 6) CD4 cell count, and 7) HIV viral load. We use Wasserstein distance (WSD) to measure the differences between real and synthetic data, which is defined as the minimum amount of probability mass that must be transported to transform one distribution into the other.

There are several notable findings. First, MeLD-generated synthetic data yield the lowest WSD for five of the seven continuous variables, while ranking second for the other two (age and CD4 cell count) (Fig. 5a, Supplementary Table 6). Second, the distribution of the total number of visits varies markedly across models, as shown by a representative synthetic dataset for each model in Fig. 5c-g. MeLD, HALO, and SynTEG all use the EOS token to indicate the final visit; however, while MeLD and HALO produce visit number distributions closely resembling those in real data, SynTEG generates data with substantially fewer visits. TimeDiff and TimeAutoDiff do not employ the EOS token but create opposite effects: the former oversamples individuals with a high number of visits, whereas the latter undersamples them. Third, while all models successfully capture the modes of CD4 cell count and HIV viral load, they struggle to represent the low-density regions observed in real data (Supplementary Fig. 1). Fourth, among MeLD variants, MeLD-Transformer demonstrates the closest fidelity to MeLD across continuous variables (Supplementary Table 6), whereas the other variants perform worse on most variables.

**Figure 5.**
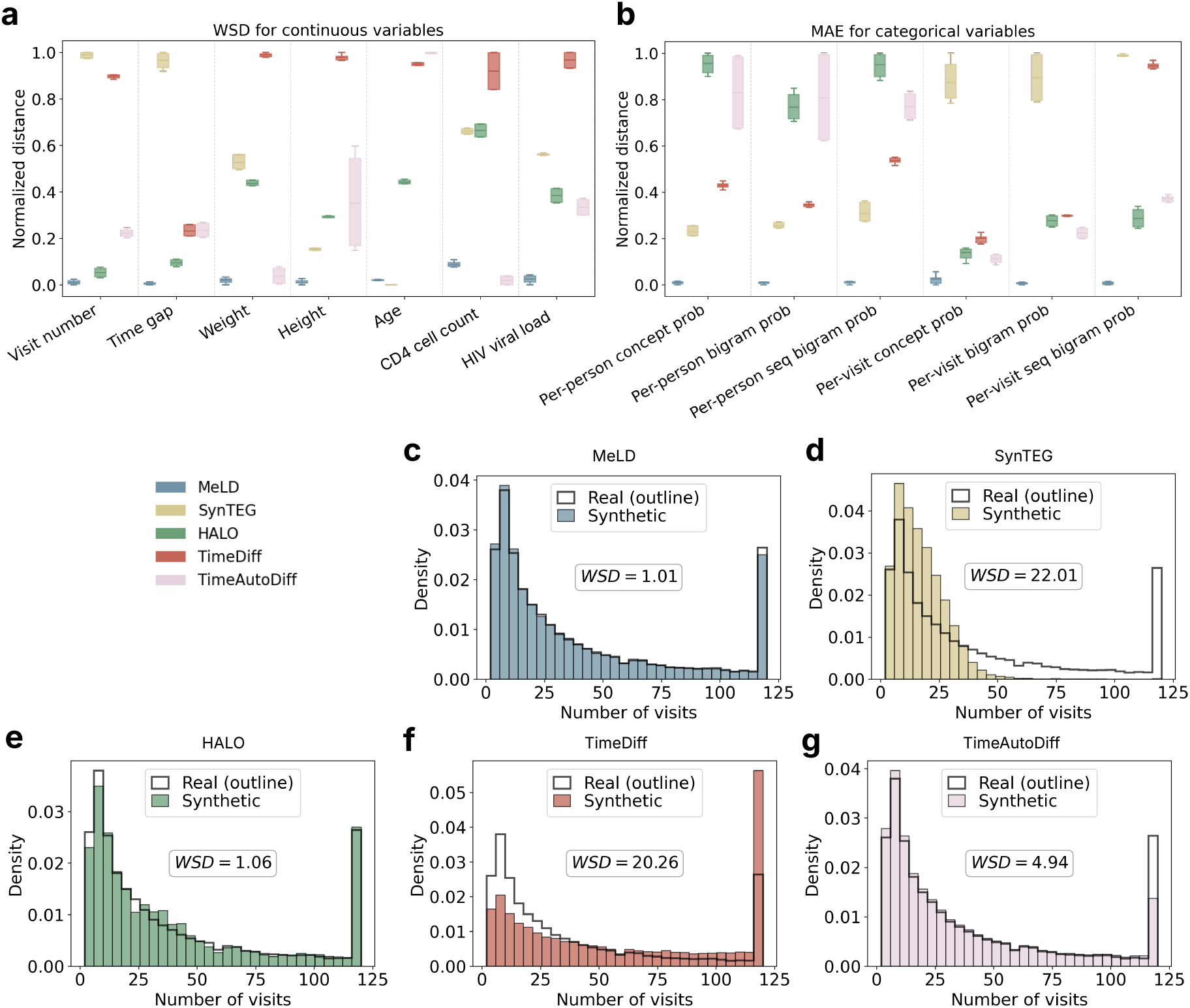
Fidelity comparison in terms of variable-level distributions. **a**, Normalized Wasserstein distance (WSD) of continuous variables, with raw values linearly mapped to the [0,1] range for each variable. **b**, Normalized mean absolute error (MAE) across categorical variables, measured using per-person and per-visit probabilities of single, bigram, and sequential bigram concepts, with raw MAE values linearly mapped to the [0,1] range. **c-g**, Probability distributions of visit numbers of real data, along with one synthetic dataset generated by each model, selected to be closest to the mean of WSD values.

For categorical variables, such as clinical diagnoses and medications, we follow the approach of Theodorou *et al*.^22^ and examine aspects at both the person- and visit-level: 1) frequencies of medical concepts (referred to as concept probability), 2) frequencies of medical concept bigrams (referred to as bigram probability), focusing on the co-occurrence of medical concepts within the same visit, and 3) frequencies of sequential medical concept bigrams (referred to as sequential bigram probability), focusing on the occurrence of medical concepts in consecutive visits. We quantify the deviation of synthetic data from real training data with respect to each of these measures using the mean absolute error (MAE).

MeLD consistently achieves the lowest normalized MAE across all measures (Fig. 5b, Supplementary Fig. 2, Supplementary Table 7), highlighting its ability to capture not only the marginal distributions of categorical variables and the co-occurrence of medical concepts within the same visit, but also their sequential occurrence across visits.

#### Correlation

Variable correlation characterizes how well synthetic data preserve the inter-variable relationships observed in the real data. We examine two types of correlations: 1) between-individual correlation, which relates individual-level means (e.g., each individual’s mean CD4 cell count value or treatment frequency) across PWH to reveal whether individuals with higher values in one variable also tend to be higher on the other, and 2) within-individual correlation, which relates visit-level deviations from each individual’s own mean (e.g., how two variables co-fluctuate around that individual’s baseline). We calculate Spearman correlation for both types across all variable pairs and report the sum of differences between synthetic and real data using Frobenius norm.

Similar model rankings are observed in between- and within-individual correlations (Fig. 6a,b, Supplementary Fig. 3), with MeLD achieving the closest correlations to real data. Although MeLD-Transformer achieves the lowest Frobenius norm distance among MeLD variants, it is outperformed by standard MeLD.

**Figure 6.**
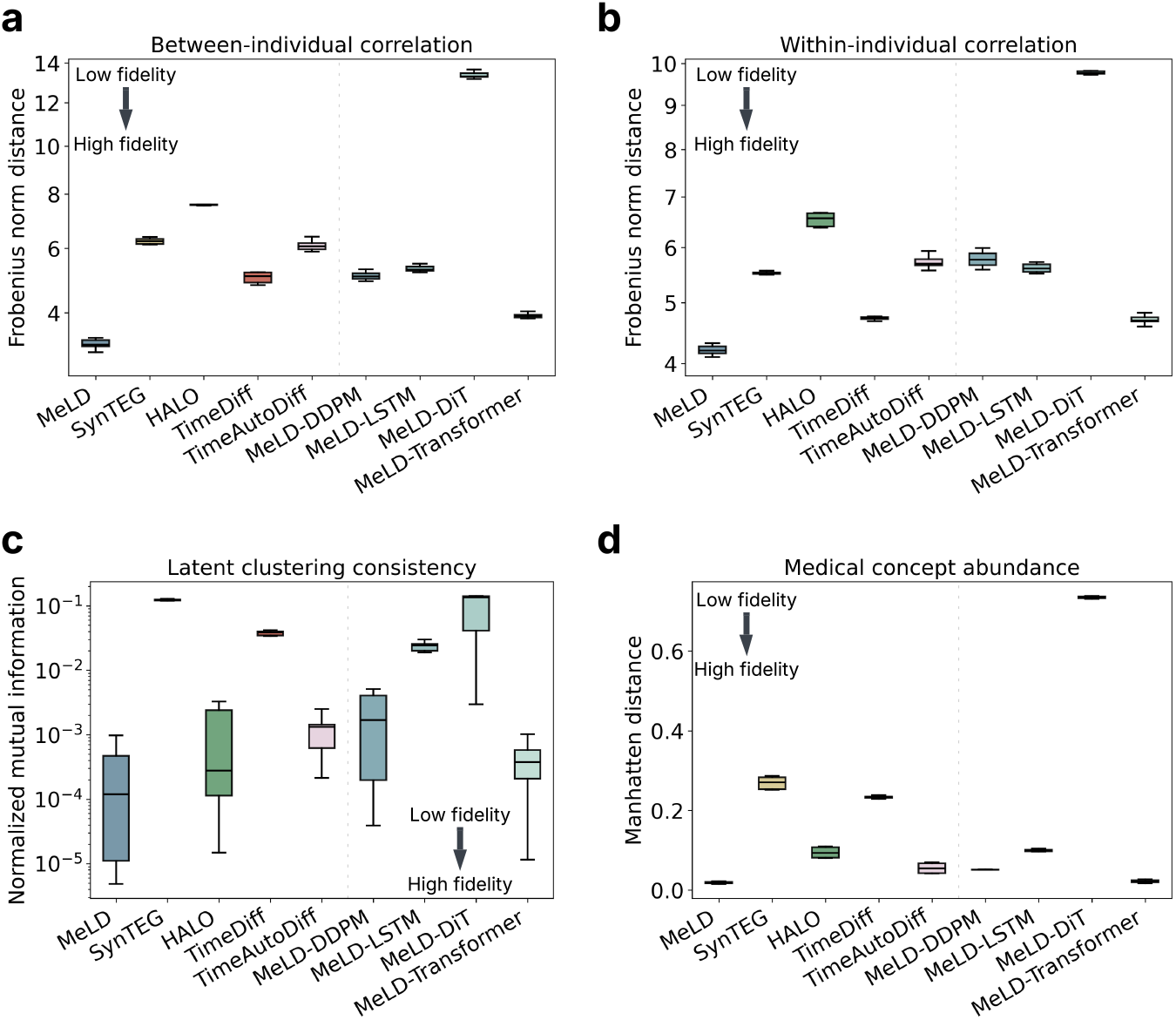
Data fidelity assessment in terms of **a**, between-individual correlation, **b**, within-individual correlation, **c**, latent clustering consistency, and **d**, medical concept abundance. Boxplots are made with ten synthetically generated datasets.

#### Latent clustering consistency

Latent clustering consistency characterizes the ability of synthetic data generation models to capture the joint distribution of all variables in real data. The procedure is conducted in the underlying latent space by 1) merging records from real and synthetic datasets, 2) applying principal component analysis and retaining components explaining 80% of the system variance, 3) conducting *K*-means clustering to identify clusters, and 4) assessing cluster-level compositions of real versus synthetic records using normalized mutual information (NMI), which measures the mutual information between cluster assignments and source labels (real versus synthetic), with the values of 0 and 1 corresponding to the best and worst fidelity of synthetic data, respectively. Synthetic data of high fidelity should produce cluster compositions that are inseparable from real data in the latent space.

The experiments show that MeLD generates synthetic data that are nearly indistinguishable from real data in the latent space (Fig. 6c, Supplementary Table 8), achieving a markedly low average NMI of 0.0003. TimeDiff and SynTEG, by contrast, exhibit the highest average NMI values of 0.0377 and 0.1117, respectively. These discrepancies are in alignment with the fidelity gap observed in other metrics that focus on the original space. Of all MeLD variants, only MeLD-Transformer matches the standard MeLD model’s performance, whereas the rest show inferior NMI.

#### Medical concept abundance

We also investigate the degree to which synthetic data generators capture the quantity of individual-level information in real data with medical concept abundance (MCA)^4^. MCA, originally designed for cross-sectional data, uses the total number of clinical events in each individual as a proxy for severity of illness or health status. We extend MCA to the longitudinal setting by aggregating visit-level MCA values and then normalizing the aggregated values with the total number of visits. The difference in the MCA distribution between real and synthetic data is then calculated using the Manhattan distance^4^.

MeLD shows the smallest MCA difference between real and synthetic data among all models, suggesting that it best preserves the overall health status distribution in real data (Fig. 6d). Moreover, the MeLD variants with different diffusion components underperform relative to MeLD, whereas MeLD-Transformer achieves a similar MCA difference.

#### Missingness pattern

Missing data are ubiquitous in observational data from healthcare records. Generating synthetic longitudinal clinical cohort data that replicate the missingness patterns observed in real data is essential to ensure realistic data quality and maintain the validity of downstream applications. Since MeLD is the only model that explicitly accounts for missingness during model training and data generation, we compare the missingness patterns between MeLD-generated synthetic data and real data prior to imputation across all continuous variables with missingness: i.e., CD4 cell count, HIV viral load, weight, and height. Missingness for each variable is characterized by two measures: 1) the overall variable-level proportion of missingness across the entire cohort (a scalar) and 2) the individual-level proportion of missingness (a distribution). It is notable that MeLD closely reproduces the proportions of missingness observed in real data, with both variable- and individual-level proportions of missingness falling within similar ranges across all variables (Table 2).

**Table 2.** Comparison of proportions of missingness between real and MeLD-generated synthetic data. Variable-level and mean individual-level proportions of missingness are reported for both real and synthetic datasets. For each metric, 95% confidence intervals are estimated over ten independently generated synthetic datasets by MeLD. Note that the definition of missingness used here differs from that used in epidemiological analyses: for any given measurement, all visits lacking an observed value are considered missing, regardless of whether the value was unmeasured, intentionally omitted, or below the detection limit.

| Variable | Variable-level proportion of missingness |  | Mean individual-level proportion of missingness |  |
| --- | --- | --- | --- | --- |
|  | Real | Synthetic | Real | Synthetic |
| CD4 cell count | 0.826 | 0.818 [0.816, 0.819] | 0.808 | 0.808 [0.806, 0.809] |
| HIV viral loads | 0.944 | 0.945 [0.944, 0.946] | 0.923 | 0.926 [0.925, 0.927] |
| Weight | 0.695 | 0.685 [0.683, 0.687] | 0.734 | 0.736 [0.734, 0.738] |
| Height | 0.947 | 0.949 [0.948, 0.949] | 0.931 | 0.936 [0.935, 0.937] |

### Privacy

To evaluate the privacy risks posed by sharing synthetically generated data, we investigate three well-studied measures: attribute inference^29,30^, membership inference^29,30^, and nearest neighbor adversarial accuracy (NNAA)^31^. For both attribute inference and membership inference, we make a reasonable assumption that the adversary has access to synthetic data but not to the underlying data generators. Real training data are used to simulate a reference scenario where this dataset is accessible to the adversary.

- Attribute inference occurs when an adversary uses synthetic data to infer unknown sensitive attributes from partial knowledge of real data. It is assumed that the adversary knows the demographic information and the 100 most common medical concepts (representing an upper bound on adversarial capability to stress-test the system) of 5,000 randomly sampled real training records. Using a K-nearest neighbor (KNN) algorithm, the most similar synthetic record among 5,000 sampled is identified to infer the value of the remaining variables. F1 score is used to quantify attack performance.
- Membership inference risk measures the extent to which an adversary can infer whether an entire real record they have access to was part of the real training data used to train the synthetic data generator. We assume that the adversary has access to a set of real records comprising 5,000 randomly sampled real training records and 5,000 randomly sampled real test records, without knowledge of their affiliation. The adversary infers that a real record was a member in the training data based on its closest distance to synthetic records, using median distance as cutoff. The proportion correctly classified is computed. A sensitivity analysis is also performed on a wide range of distance cutoffs.
- NNAA risk measures the degree of overfitting of a synthetic data generator to the real training data, using a held-out real dataset (e.g., real test set) as a reference. When a synthetic data generator inappropriately memorizes real training data rather than generating novel samples, the generated synthetic records tend to be systematically closer to real training data than to the held-out real test data. To measure NNAA risk, we randomly sample 5,000 records each from the real training, real test, and synthetic datasets, and compare the aggregated minimum distance of each synthetic record to their nearest neighbor in the real training and test sets, and vise-versa. A positive NNAA value indicates that synthetic data are closer to the training data and vise-versa. We use a threshold of 0.03 to determine model overfitting, as suggested by Yale *et al*.^31^.

Fig. 7 depicts the three privacy risk metrics for the synthetic data generated by MeLD and the baseline models, and reveals several notable findings. First, attribute inference risks, characterized by F1 scores, range from 0.020 to 0.050 across all models, suggesting that attribute inference attacks in general are ineffective for both MeLD and other synthetic data generators. MeLD ranks in the middle, and its variants exhibit comparable levels of risk (Supplementary Table 9). Second, except for HALO, membership inference risks are nearly the same for all models, achieving an accuracy around 0.510, which is similar to random guessing. A sensitivity analysis across a wide range of distance cutoffs produces consistent observations (Supplementary Fig. 4). Third, MeLD attains an average NNAA risk of -0.0008, substantially below 0.03 threshold, suggesting minimal model overfitting. In contrast, such risk is elevated for TimeDiff (0.031 [0.012, 0.050]), a level that can cause privacy leakage. In summary, MeLD and its variants maintain strong privacy protection, which implies that improved utility and fidelity are not achieved at the expense of privacy.

**Figure 7.**
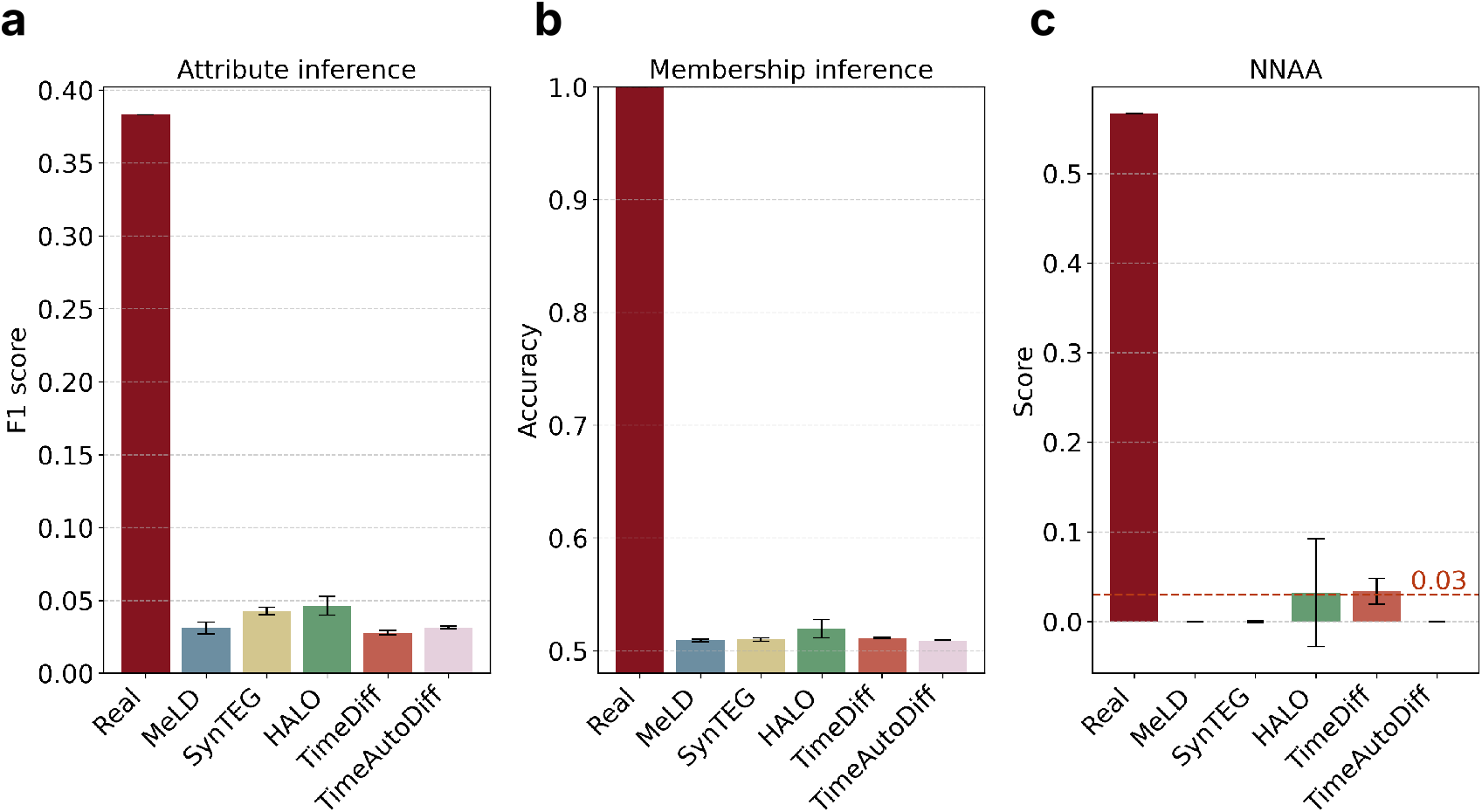
Privacy risk evaluation. **a**, F1 score of attribute inference attack. **b**, accuracy of membership inference attack. **c**, NNAA risk. The risks associated with real data are shown in the leftmost red bars. NNAA risk less than 0.03 is deemed acceptable. 95% confidence intervals are estimated over ten synthetically generated datasets. NNAA=nearest neighbor adversarial accuracy.

## Discussion

Generating long synthetic data sequences has been a challenge across a variety of domains, particularly in time-series modeling that contains a large number of mixed-type variables^32^. MeLD addresses this issue through a latent diffusion-based framework integrated with a transformer backbone, which enables the generation of long-term health trajectories with realistic longitudinal dependencies. Our empirical investigation demonstrates that MeLD substantially outperforms state-of-the-art methods designed for longitudinal EHR data generation across evaluation dimensions (Fig. 1e). Notably, MeLD achieved the best fidelity in both continuous and categorical variables, highlighting its flexibility in handling mixed-type data. MeLD also exhibited the strongest consistency in capturing both within- and between-individual correlations. Moreover, the close alignment in latent-space structure and medical concept abundance reaffirms that MeLD effectively generates synthetic data following the joint distribution of the real data. Importantly, MeLD is also designed to natively handle missingness in cohort data, a fundamental characteristic of real-world observational data that most previous studies have overlooked. We attribute these encouraging results to the versatility of the VAE architecture and the exceptional generative capabilities of the diffusion transformer. Specifically, our VAE’s encoder design embeds complex data patterns into a unified latent representation, allowing the diffusion module to learn and generate data without directly dealing with the complexity of the original feature space. Meanwhile, the transformer-based diffusion mechanism allows MeLD to effectively capture global population structures and generates detailed temporal and cross-sectional patterns, resulting in synthetic cohorts that closely mirror real-world individual variability.

Our focus on assessing the utility of synthetic longitudinal cohort data through time-to-event analyses and risk-factor estimation represents a largely unexplored yet critical dimension of synthetic health data research. Unlike most prior studies that produce general-purpose synthetic data and narrowly assess data utility for predictive modeling or data augmentation, our research explicitly examines synthetic data capability to support inferential tasks that are central to clinical and epidemiological research. This focus closely aligns the broader vision of leveraging synthetic data to enable scientific hypothesis generation. While one cannot enumerate all potential hypotheses for utility evaluation, evaluating their performance through representative inferential or replication-oriented tasks provides a practical and rigorous means to estimate their real-world research value.

Prior evidence suggests that synthetic data tend to produce unacceptably high error rates, particularly false-positive findings (type I errors)^33^. In contrast, our evaluation demonstrates that MeLD commits only a single error across 40 tested scenarios, generally preserving the underlying statistical relationships. This type of evaluation, as well as the encouraging results, establish an empirical foundation for considering synthetic longitudinal data as a credible bridge toward exploratory analysis and hypothesis-driven biomedical research.

Throughout the experimental results, we observed that architectural choices play a critical role in preserving long-range temporal coherence. Even when using real clinical visit sequences containing up to 120 visits and spanning more than three decades, MeLD demonstrated advantages over state-of-the-art designs in capturing long-range dependencies. Further notable, replacing a GRU with a transformer in the VAE component of MeLD did not produce a clear performance advantage. This observation likely reflects the functional division within the latent diffusion design, where VAE’s RNN is responsible for learning temporal dependency between visits and aligning them closely in latent space^34^, whereas the transformer in the diffusion component captures higher-order temporal structure and drives generative dynamics. As a result, a lightweight GRU fulfills the VAE’s role in this design.

While MeLD demonstrates the strongest overall performance, some metrics still favor specific existing models. For example, TimeDiff, an LSTM-based diffusion model, achieves the lowest FDR in time-to-diagnosis analyses. This advantage likely reflects the short time-to-disease durations in the real cohort of PWH. In such conditions, recurrent architectures like LSTM may better capture short-term temporal dependencies. However, its advantage diminishes for outcomes requiring modeling extended temporal horizons, such as maximum follow-up time and time-to-death, where MeLD demonstrates superior robustness. Similarly, HALO and SynTEG outperform MeLD in capturing the tails of continuous variable distributions (Supplementary Fig. 1). This difference arises because both models apply post-hoc sampling from the discretized empirical distributions for reconstruction, whereas MeLD reconstructs data directly from a compressed latent representation. Consequently, MeLD follows the VAE’s tendency to emphasize high-probability regions of the data space, leading to slightly reduced tail coverage. Nonetheless, this mechanism allows MeLD to achieve higher fidelity in overall distribution and inter-variable correlations.

The tradeoff between data utility and privacy in machine learning-based synthetic health data generation has been reported in multiple prior studies, where higher utility is generally linked to lower privacy protection (or higher risks), and vice versa^3,4^. In these works, “utility” is typically defined in terms of fidelity to real data and performance on downstream tasks. Most of these observations are derived from synthetic snapshot or cross-sectional health tabular data. However, we do not observe such a phenomenon in the scenario of synthetic longitudinal clinical cohort generation. Specifically, MeLD, the generator achieving the highest performance in nearly all utility and fidelity metrics, also exhibits low privacy risks, compared to state-of-the-art models. This observation implies that the conventional utility-privacy Pareto frontier observed in prior research may not apply to all modeling scenarios. Rather, it reflects the limited representational capability of previous models to learn complex longitudinal structures without memorizing individual data patterns. MeLD’s architectural design may allow the model to capture high-level regularities from real data while reducing the likelihood of reproducing real data. As a result, we believe the existing frontier between utility and privacy is not necessarily optimal in synthetic longitudinal cohort generation, and substantial opportunities remain for innovation in model design to enhance data utility and fidelity without sacrificing privacy. We are releasing MeLD as an open-source tool, so that it can enable HIV researchers, and epidemiologists more broadly, conducting cohort studies to generate and share synthetic datasets to facilitate research replication and transparency. We believe MeLD bridges the long-standing divide between data protection and utility, and paves the way for a more inclusive, collaborative, and reproducible era of global HIV and epidemiological research. Beyond a methodological advance, we are releasing the synthetic cohort of PWH generated by MeLD, which is composed of over 49,000 simulated PWH. This dataset represents the first high-quality synthetic longitudinal cohort of PWH that mirrors the structural and temporal complexity of data typically seen in large international HIV consortia. This will enable researchers with diverse expertise backgrounds, particularly those without access to proprietary cohort data, to actively engage in hypothesis generation, methodological innovation, and model benchmarking, thereby accelerating the advancement of HIV research and treatment. This dataset can also be used in education and training in HIV medicine by allowing trainees to interact with and explore real-world-like datasets. We note that Kuo *et al*.^35^ also released a synthetic cohort of PWH, but that their dataset contains substantially fewer variables and limited clinical depth (15 variables) compared to MeLD.

While our results suggest that MeLD is a major step forward in longitudinal synthetic data generation, there are opportunities for future improvement. First, the evaluation of MeLD is conducted using a single longitudinal dataset, which may limit the generalizability of our findings. Broader evaluations of MeLD across additional chronic diseases and data settings will be pursued in future studies. Nevertheless, the CCASAnet cohort, one of the largest and most diverse international HIV datasets, offers extensive population heterogeneity and a harmonized data structure, which enhance the external relevance of our results to other HIV consortia. Second, our evaluation includes four state-of-the-art models but does not cover all existing methods for longitudinal data generation, such as TarDiff^36^, FourierFlow^37^, DiffusionTS^38^, TimeGAN^39^, and TimeVAE^40^. TarDiff is tailored for generating synthetic longitudinal health data that are particularly helpful in improving fairness in specific downstream prediction tasks, while FourierFlow and DiffusionTS target univariate time-series generation, making them less suitable for our objective of modeling multivariate clinical trajectories. TimeGAN and TimeVAE, although well-known, are not considered state-of-the-art and have been shown to perform less effectively than the baseline models we select^8^. Our selected baseline models encompass the primary generative AI paradigms for general-purpose synthetic longitudinal data generation, providing a representative foundation for our evaluation. Lastly, synthetic data might introduce bias and fairness issues that affect data quality across subgroups. This study does not perform group-specific evaluations and MeLD is trained without conditional guidance, which could affect subgroup fidelity. Addressing these issues represents an important avenue for future research.

## Methods

### Data

This study uses the PWH cohort data from the Caribbean, Central and South America Network for HIV Epidemiology (CCASAnet)^41^, comprising information collected from HIV clinical sites in seven countries: Centro Medico Huesped, Buenos Aires, Argentina (Argentina-CMH); Instituto de Pesquisa Clinica Evandro Chagas, Fundação Oswaldo Cruz, Rio de Janeiro, Brazil (Brazil-FC); Fundacion Arriaran, Santiago, Chile (Chile-FA); Les Centres GHESKIO, Port-au-Prince, Haiti (Haiti-GHESKIO); Instituto Hondureño de Seguridad Social and Hospital Escuela, Tegucigalpa, Honduras (Honduras-IHSS/HE); Instituto Nacional de Ciencias Médicas y Nutrición Salvador Zubirán, Mexico City, Mexico (Mexico-INCMNSZ); and Instituto de Medicina Tropical Alexander von Humboldt, Lima, Perú (Peru-IMTAvH). Clinical data were collected at each site, de-identified, and sent to the CCASAnet Data Coordinating Center at Vanderbilt University (Nashville, TN, USA) for harmonization and processing. This study does not include data from Peru because its approval period differs from that of the other sites. Institutional ethics review boards from all other sites and Vanderbilt University reviewed and approved the project. All PWH of at least 18 years old at the time point of program enrollment, who have at least 2 visits are included in this study. Descriptive statistics of the dataset are shown in Supplementary Table 1, and the variable list can be found in Supplementary Table 2.

Given the study population 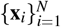, we adopt the standard notation for longitudinal EHR data, where the record of individual *i* with *t*_*i*_ historical visits is denoted as 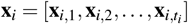. Each visit shares the same set of *J* variables, and the trajectory of variable *j* across all visits is represented as **x**_*i*,; *j*_. It is important to note that the original clinical cohort data are typically 1) organized in a set of relational database tables and linked through unique identifiers, 2) longitudinal with variable-length follow-up: each individual’s record 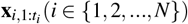 is a sequence of visits of length *t*_*i*_ *T*, where *T* represents the maximum number of visits across the cohort, and 3) of mixed-type: each visit 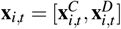 consists of *s continuous* variables 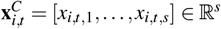 and *k binary* variables 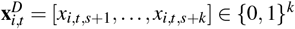, which can be derived from categorical or discrete variables.

To preprocess the original CCASAnet data, we integrate visit information of medical events from distinct tables, as well as corresponding timestamps, into unified clinical entries. More concretely, laboratory measurement results (e.g., CD4 cell count and HIV viral load) and medical concepts (e.g., clinical diagnoses and treatment regimens) are integrated into visit-level representations, ensuring that all relevant information for each visit is consolidated into a single clinical entry. Inter-visit time gaps are added as a continuous variable to each visit. All continuous variables are normalized to [0, 1] in a linear manner, whereas categorical variables are encoded as one-hot representations. Inspired by HALO^22^, demographic information (i.e., age, sex, and site) is indicated in “visit 0”. These processing steps yield a complete clinical visit sequence 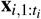, where each entry contains a snapshot of the individual’s health status corresponding to a specific clinical visit. To manage computational efficiency, we limit each individual’s record to a maximum of *T* =120 visits, which corresponds to the 90th percentile of visit counts across the cohort. The resulting preprocessed dataset contains 135 variables and an average of 45 visits per person, spanning over three decades.

To enable batch processing of variable visit lengths, we use 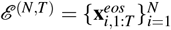 to denote the end-of-sequence (EOS) tokens, where 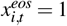 if visit *t* is the last visit (i.e., *t* = *t*_*i*_), and 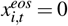 otherwise. Moreover, we introduce 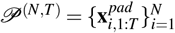 to denote the masking tokens used to mark the padding positions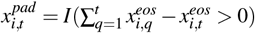. This enables implementing zero-padding to construct a uniform-length data matrix 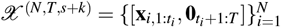 In addition, the timestamps of all individuals’ records are represented as time tokens 𝒯 ^(*N,T*)^.

To explicitly handle missing data in relevant continuous variables, whether due to uncollected values during clinical visits or laboratory results falling below detection limits, we use 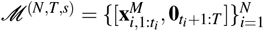 as the missingness indicators to mark missing entries, where 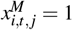 indicates a missing value, and 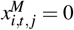 otherwise. MeLD leverages these tokens within the input data to capture missingness patterns while masking out missing values so that model training focuses on the observed values. In the post-processing stage, we use the learned missingness indicators to determine the positions of missing values. For a fair comparison with models that do not natively handle missing values, missing values are imputed with Multiple Imputation by Chained Equations (MICE)^25^ before training.

### Model architecture

As illustrated in Fig. 1, MeLD contains two major components: 1) a variational auto-encoder (VAE) that learns a mapping between the original sparse, mixed-type data space and a dense, continuous latent space, and 2) a transformer-based diffusion model to generate synthetic longitudinal visits in the latent space.

1. **VAE:** Variables of each data type stream are first embedded through the corresponding embedding layer. An MLP layer then takes the concatenation of embeddings from all variable type streams and fuses them into a unified embedding. The fused embedding, sized (*N, T, H*), is reshaped into (*NT, H, B*) using a 1D convolution layer with *B* channels. This flattened representation is then passed into a transformer encoder to learn the dependencies among variables, with the output being reshaped back to (*N, T, H*) via another 1D convolution layer. Next, the output, together with MLP-embedded real timestamps and position-encoded visit indices, are passed into two RNN modules in parallel to learn temporal dependency, yielding *μ*_*z*_ and 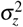 as the mean and variance of the approximate posterior distribution over the latent variables. The decoder takes the synthetically generated visit sequence in the latent space, sized (*N, T, L*), as input, transforms it using an MLP, and finally applies type-specific linear projections to map them back to the original spaces. This architecture follows the TimeAutoDiff VAE in design, but largely enhances its capability and flexibility by incorporating the EOS signals and missingness so that MeLD can more accurately replicate follow-up complexity and missingness patterns in longitudinal clinical cohorts.
2. **Transformer-based diffusion:** We adapt DiT^19^, a transformer-based diffusion model designed for image generation, to the task of clinical visit sequence generation. Specifically, data sampled from the latent space learned by the VAE encoder are used as input with dimension (*N, T, L*), and are iteratively corrupted with Gaussian noises during the forward process of diffusion. Corrupted latent visit sequences are then leveraged to iteratively remove the added noises and finally recover the underlying latent visit sequences. In particular, at each reverse step, input data are first transformed via a 1D convolution layer for patchification and passed to a stack of DiT modules for dependency modeling and noise prediction. In our design, we treat each visit as an individual patch for convenience. Within the DiT module, the patched sequences passes through normalization, attention, and point-wise feedforward layers. In addition, the timesteps of the diffusion process serve as a conditional input for DiT, which are first embedded with an MLP layer and then applied to scaling and shifting transformation before and after the attention and the feed-forward layers in DiT.

Beyond the aforementioned overall design, MeLD introduces multiple architectural innovations compared to baseline models. First, unlike the TimeAutoDiff VAE, which creates a separate embedding for each variable, an approach that scales poorly with a large number of variables, we employ a more efficient strategy that uses a single embedding module per variable type and aggregates visit-level embeddings within each type. Second, we do not consider additional longitudinal modules in the decoder, as informed by the findings observed by Suh *et al*.^23^ that such design does not bring benefits to overall data quality. Li *et al*.^42^ also show that strong decoders can undermine longitudinal information in latent representations, thereby leading to VAE posterior collapse and degraded generative performance in diffusion models. Third, we utilize a diffusion training paradigm that is optimized for faster convergence rate and better data quality (i.e., FasterDiT) compared to the standard version^20^.

We follow the two-stage training strategy of latent diffusion models^34^, where we train the VAE in the first stage, and the diffusion model in the second. To generate synthetic longitudinal records with the trained MeLD, we first sample a visit sequence in the latent space from the learned diffusion model and then decode it through the VAE decoder to obtain the synthetic record in the original data space. More concretely, the model input **X** = {𝒳, *𝒯, ℰ, 𝒫, ℳ }*, comprising real data matrix, timestamps, EOS tokens, padding masks, and missingness indicators, is fed into the VAE encoder **Enc**(· ) to derive the mean *μ*_*z*_ and variance 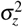 of the latent posterior distribution *p*(**Z**|**X**). For the reconstruction task, variational samples drawn from this posterior, **z** ∼*p*(**Z**| **X**), are provided to the decoder **Dec**(· ) and diffusion for their training. This architecture forces the encoder to learn semantically rich latent representations that capture temporal dynamics, variable dependencies, sequence lengths, and missingness patterns. As a result, latent samples generated by the diffusion model inherently encode the information necessary to generate mixed-type, variable-length longitudinal cohort data.

During data generation, novel latent samples **z**^′^ are first produced by the diffusion model and then provided to the VAE decoder, which generates synthetic data matrix 𝒳 ^′^, EOS tokens ℰ ^′^, and missingness indicators ℳ ^′^. The synthetic data are subsequently reconstructed into a meaningful visit-level representation by applying ℰ ^′^ and ℳ ^′^ to 𝒳 ^′^. This process yields tabular data in which each row represents a clinical visit of an individual, accommodating variable-length follow-ups, mixed-type variables, and missing values. For evaluation, we post-process the MeLD-generated data by imputing missing values with MICE, following the same preprocessing procedure applied to the real data. In addition, we retain an unimputed version to evaluate the missingness patterns in the synthetic data.

The first stage trains a VAE with a 256-dimensional hidden layer, two-layer GRU and MLP modules, and an eight-head transformer with 64 channels. It is optimized with a batch size of 128 to yield latent representations with 32-dimensional latent size. The second stage trains a transformer-based diffusion with a 256-dimensional hidden size and eight layers of eight-head attention, using a batch size of 1,024.

#### VAE loss

The objective function of VAE minimizes the combined loss of the data reconstruction term, ℒ_*RE*_, and the KL-divergence term, *ℒ*_*KL*_, which measures the divergence between the posterior distribution *p*(**Z**|**X**) and the prior *p*(**Z**):

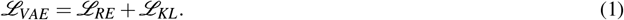

### Reconstruction Loss

Due to the mixed-type, variable-length nature of clinical cohort data, different loss functions are needed: we use mean square error (MSE) normalized by time for continuous variables, and cross-entropy loss normalized by time for categorical variables and special tokens.

First, taking special tokens into account, we calculate the reconstruction loss for **continuous variables** as:

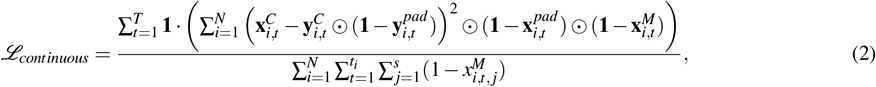

where **y** represents the reconstructed data and ⊙ denotes the element-wise multiplication operation. This design excludes imputed values from the loss calculation to maintain the model’s robustness to imputation quality.

Second, we calculate the reconstruction loss for **categorical variables** as:

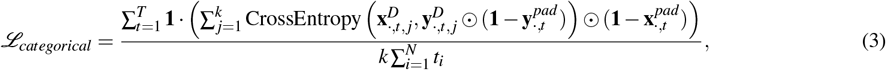

where we denote a visit vector containing all individuals as **x**_;*t*_.

Next, we calculate the reconstruction loss for **missingness indicators** (treated as categorical variables) as:

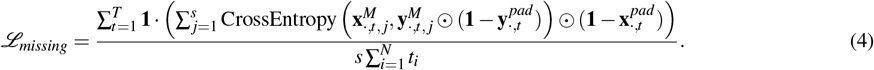

Finally, we calculate reconstruction loss for the **special EOS token** as:

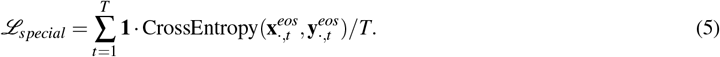

This loss is essential for enabling the model to learn the semantics of EOS and padding tokens and to generate records with variable lengths.

The total reconstruction loss is the sum of the aforementioned four losses:

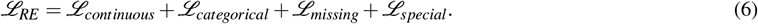

### KL-divergence Loss

The KL-divergence loss is

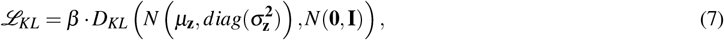

which is to minimize the divergence between the posterior, defined by *N μ*_*z*_, *diag*(*σ*_*z*_^2^), and the Gaussian prior *N*(**0, I**). We apply a weighting factor *β* to achieve a balance of smooth concentrated posterior function while maintaining its expressiveness.

Such a mechanism is crucial to prevent posterior collapse (a phenomenon in which the posterior distribution is forced to a standard Gaussian), thereby avoiding a trivial Gaussian diffusion model^42–44^.

#### Diffusion loss

The VAE encoder reduces mixed-type variable-length clinical cohort data into a fixed-dimension continuous vector, which enables us to employ a simpler design for the diffusion transformer similar to the ones used for image generation^19^. Given the latent encodings **z**_0_ and Gaussian noise *ε* ∼*N*(**0**, *I*), the forward diffusion process defines a sequence of progressively noisier samples as

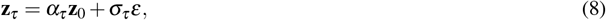

where *α*_*τ*_ and *σ*_*τ*_ are functions of the diffusion timestep *τ* ∈ (0, ∞). *σ*_*τ*_ increases with *τ*, whereas *α*_*τ*_ decreases. As a result, the data distribution gradually approaches an isotropic Gaussian as *τ* → ∞. The corresponding reverse process learns to iteratively denoise samples by estimating the noise *ε*_*θ*_ (**z**_*τ*_, *τ*) at each timestep. A diffusion model can be trained with a simple MSE loss between added noise and the predicted noise, which allows the accurate reconstruction of the target distribution *p*(**z**_0_) during sampling^24^:

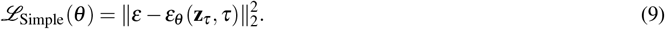

Alternatively, the reverse process can be formulated as a probability flow ordinary differential equation (ODE) that transforms a Gaussian noise to the target distribution *p*(**z**_0_)^45,46^. In this formulation, the ODE is governed by a conditional velocity field 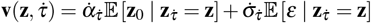], where 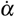] and 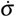] are functions of 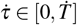] . It represents the expected direction and rate at which a noisy sample **z** should change at time 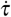] in order to move toward the target distribution *p*(**z**_0_). This quantity can be estimated using a neural network 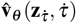]. Thus, a diffusion model can also be trained with the flow matching objective:

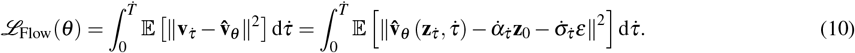

Empirical evidence^20^ has shown that Eq. 10 achieves superior performance and higher sampling efficiency than Eq. 9. Yao *et al*.^20^ propose a linear schedule 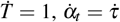 and 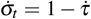 for the ODE strategy and develop FasterDiT. In addition, FasterDiT uses cosine regularization to encourage similar samples to cluster in hidden space, a strategy conceptually aligned with the regularization proposed by Wang *et al*.^47^. Taken together, the objective function used for tuning MeLD’s diffusion model is:

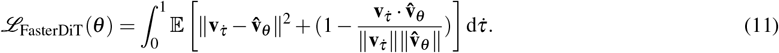

### Evaluation framework

To account for randomness in model training, we train each model independently twice. Each trained model then generates five synthetic datasets. Thus, a total of ten datasets are used for evaluating the underlying model’s capability. These ten datasets, referred to hereafter as replicates, are generated using distinct random seeds and maintain the same sample size as the real training data. For all evaluation metrics, we report the mean and 95% CIs across replicates. The 95% CIs are constructed with normal approximation, with the mean and standard deviations of ten replicates serving as the estimates of the sampling distribution’s parameters. To ensure fair comparison among models that cannot directly handle missing data, missing values in real data are imputed prior to evaluation. We now describe the evaluation metrics used to assess data utility, fidelity, and privacy.

#### Time-to-event estimation

All medical events and their corresponding timestamps are extracted for each individual. For a given event, e.g., a diagnosis of a certain disease (clinical endpoint), if it does not occur in an individual’s medical history, then this individual is treated as censored for this clinical endpoint, with the time of the last recorded visit used as the censoring time. We calculate both Kaplan-Meier and Nelson-Aalen estimates: the Kaplan-Meier estimator is used for quantifying differences between survival curves, whereas the Nelson-Aalen estimator is chosen for visualization. Two Kaplan-Meier-based methods are used to evaluate differences between survival curves. The log-rank test compares the observed and expected numbers of events between groups at each event time, and aggregates these differences across all event times to assess whether survival curves differ, assuming equal survival under the null hypothesis. The log-rank test is widely used to compare survival distributions between groups. However, it has limited power when survival curves cross or when differences occur late in follow-up, as the number of individuals at risk diminishes. To complement the log-rank test, we compute the non-parametric Kaplan-Meier distance (KM-D)^26^, calculated as the mean absolute difference between two Kaplan-Meier curve estimates evaluated at 1,000 equally spaced time points from time 0 to the last observed event in the shorter curve.

In addition, we evaluate the maximum follow-up time *T*_*max*_ in a cohort as a supplement metric to show if synthetically generated data fail to reproduce the maximum observation time in real data.

Time-to-diagnosis analysis focuses on the time from ART initiation to the first recorded diagnosis of a clinical endpoint. We perform the log-rank tests across all 50 clinical endpoints and apply a Bonferroni correction to adjust for multiple tests, which yields an adjusted significance threshold of 0.0001. We then report the overall false discovery rate (FDR), as well as the mean of KM-D calculated from 50 clinical endpoints over ten replicates.

#### Prediction

An important use case of synthetic clinical cohort data is to support the development of outcome prediction models. We select three representative disease prediction tasks and use data observed during the first 180 days since program enrollment as predictors to predict whether an individual will subsequently develop the disease. Individuals are excluded if they were diagnosed within the first 180 days or their total follow-up time is shorter than 180 days. In data preprocessing, PWH from Haiti are excluded from analysis as relevant clinical endpoints are unavailable. All available variables, including demographics, weight, height, CD4 cell count, HIV viral load, ART regimens, and clinical diagnoses, are utilized as predictors. A two-layer GRU model with 256 hidden units is trained, with a 9:1 random training-validation split. Early stopping with a patience of five epochs is utilized to determine the end point of model training. Model performance is evaluated using AUROC on real test dataset, comparing the reference model trained on real training data against models trained on synthetic datasets.

#### Risk factor estimation

Risk factor estimation assesses how well synthetic data reproduce the clinically meaningful associations observed in real data. We first construct the survival dataset based on the longitudinal cohort data of PWH by defining the time-to-event and censoring indicators and identifying the baseline risk factors, including sex, age, CD4 cell count, and calendar year at ART initiation. Then the Cox-proportional hazard model is used to estimate hazard ratios, following model specifications consistent with published epidemiology research using CCASAnet data^28,48^. Estimates obtained from models fitted on real training data serve as the reference for comparison.

While there is no consensus on how to evaluate data utility in risk factor estimation, we propose a set of complementary metrics for this purpose. First, with the statistical significance set at 0.05 and the estimates from real data as the “ground-truth”, we count type I and type II errors in inferences derived from synthetic data. We also calculate the number of direction errors, which happen when the signs of statistically significant effects identified from the synthetic data are in the opposite direction to those found in real data. Together, these errors measure how consistently analyses performed on synthetic data would lead to the same conclusions in risk factor effects as those based on real data. Second, we compare the pooled effect estimates for each risk factor using Rubin’s rule by combining the means and standard error estimates across ten synthetic replicates. This evaluation is used to assess the magnitude of discrepancies between estimated effects based on real and synthetic data.

To dive deeper into the numerical differences in the estimates, we also report the following metrics in Supplementary Table 5: 1) bias, averaged over the ten replicates, which quantifies the deviation between estimates from the synthetic data and those from real data, i.e., *β*_*syn*_ −*β*_*real*_, 2) standard error (SE) ratio, calculated as the ratio of the SE derived from synthetic data to that from real data, averaged over the ten replicates, i.e., *se*_*syn*_*/se*_*real*_, and 3) CI coverage, calculated as the fraction of the real CI that is covered by the CI derived from synthetic data, normalized by the total span of their union to account for potentially wider synthetic CIs. This metric assesses both the estimate deviations and their uncertainty.

#### Variable-level distributions

To quantify variable-level fidelity, we calculate the distances between real and synthetic distributions for both continuous and categorical variables. For each continuous variable, we calculate the Wasserstein distance (WSD) between its marginal distribution in real training data and in each synthetic data replicate. WSD measures the minimum amount of probability mass that must be transported to transform one distribution into another. We report the mean WSD across all replicates along with the corresponding 95% CI.

For categorical variables, we evaluate the frequencies of individual variables (i.e., medical concepts encompassing diagnoses and treatment regimens), their bigrams (i.e., co-occurrences within the same visits), and sequential bigrams, representing pairs of concepts observed in two consecutive visits. The evaluations are conducted at both the person and visit levels. In the person-level analysis, multiple occurrences of the same medical concept or concept pair across an individual’s visits are counted only once, whereas in the visit-level analysis, each occurrence of a concept or concept pair is counted separately. Specifically, concept probability focuses on the occurrence of individual medical concepts, while bigram probability captures how often two distinct medical concepts co-occur within the same visit. In contrast, sequential bigram probability measures the frequency with which a concept pair appears across consecutive visits.

For both person- and visit-level analyses, we then derive the aggregate differences between the real training and synthetic datasets using mean absolute error (MAE), which characterizes the magnitude of the differences. We report the means and corresponding 95% CI across ten replicates.

#### Correlation

While correlation is a standard metric for evaluating synthetic cross-sectional data fidelity, it has rarely been applied in the longitudinal data setting. To address this issue, we propose to evaluate two types of correlations: between-individual and within-individual correlations. For the between-individual correlation, we first compute the mean of each variable across all visits for each individual 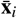, and then derive variable-variable correlations using Spearman’s rank correlation coefficients, chosen because of the mixed data types. For the within-individual correlation, we first center each individual’s variable values using the corresponding variable-level mean across all visits 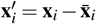 and then calculate variable-variable correlations on the centered data matrix **X**^′^ using the same Spearman correlation method. For both correlation types, the absolute differences of the correlation matrices calculated from the real training and synthetic datasets are aggregated using Frobenius norm to provide an overall measure of the discrepancy. We report the mean of Frobenius norm distances across ten replicates, as well as the corresponding 95% CI.

#### Latent clustering consistency

We employ latent clustering analysis to assess how effectively a synthetic data generation model captures the joint distribution of real data in the latent space. At a high level, we perform data clustering over the combined set of real and synthetic data and then investigate the composition of each resulting cluster to assess how well the two data sources are intermixed.

We reshape the 3D data matrix of size (N, T, s+k) into a 2D matrix of size (N,T× (s+k)), where each row represents the complete record of an individual. For computational efficiency, 5,000 records are randomly selected from both the real training and synthetic datasets, resulting in a combined total of 10,000 records. We then perform principal component analysis (PCA) to the merged datasets, and select the principal components that explain 80% of total variance to project the data from the original space into a latent space. *K*-means clustering is then applied across a range of clusters number *K* from 2 to 10, and the optimal *K* is selected as the one that leads to the smallest silhouette distance. With all clusters identified, normalized mutual information (NMI) is calculated to measure the agreement between two clustering assignments: the cluster labels produced by *K*-means versus the true labels (real versus synthetic):

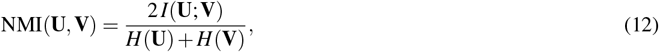

where *I*(**U**; **V**) is the mutual information between partitions **U** and **V**, and *H*(·) denotes entropy. We report the mean NMI and the corresponding 95% CI calculated from ten replicates.

#### Medical concept abundance

The above data fidelity metrics all evaluate variables and their interrelationships. To examine whether information at the individual level is also well preserved, we measure the medical concept abundance (MCA)^4^, a metric originally designed for assessing synthetic cross-sectional health data. MCA serves as a proxy of a population’s distribution of illness burden or health status, which enables evaluating how faithfully synthetic data reflect real individual-level health distributions. In the cross-sectional context, MCA for an individual is computed by counting the number of distinct medical concepts documented in their record:

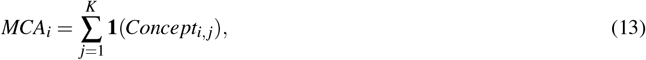

where *K* denotes the size of the medical concept space. To quantify the difference in MCA between real and synthetic data, the empirical MCA distributions built from the real and synthetic datasets are first discretized to a fixed number of bins and then normalized. The Manhattan distance of the two discretized distributions is reported.

To extend the original MCA metric to longitudinal clinical records with variable visit sequence lengths, we calculate the total occurrences of medical concepts within individual *i*’s longitudinal record and then normalize it by the number of visits *t*_*i*_:

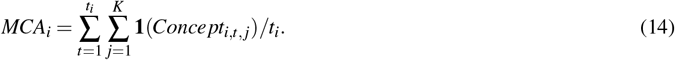

In this study, we consider all clinical diagnoses and treatment events as medical concepts. The same distance measure described above is then applied to quantify the longitudinal MCA difference between the real and synthetic datasets. We report the mean distance of MCA and the corresponding 95% CI calculated from ten replicates.

#### Missingness pattern

The definition of data missingness varies across disciplines and applications. In this study, for evaluation purposes, we treat all unobserved measurement values in each visit (i.e., CD4 cell count, HIV viral load, weight, and height) as missing data, whether they are unmeasured, intentionally omitted, or below the detection limits. With 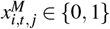 defined as the missingness indicator for individual *i*, visit *t* and variable *j*, two measures are considered: 1) variable-level proportion of missingness, defined as the overall missingness proportion for variable *j*, 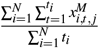, and 2) individual-level proportion of missingness, defined as the missingness proportion for variable *j* within an individual, 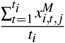. We report the mean and 95% CI of both proportions in synthetic data replicates and the real training dataset.

#### Privacy

Privacy evaluation focuses on three types of risks: attribution inference, membership inference, and nearest neighbor adversarial accuracy (NNAA). Following the approach of Theodorou *et al*.^22^, all continuous variables are discretized into 30 quantile-based bins, which converts the entire dataset into a representation of binary medical concepts to facilitate consistent risk assessment. And for each experiment, we report the mean and corresponding 95% CI calculated from ten replicates.

For attribute inference attacks, an adversary is assumed to possess partial knowledge of real individual’ records, and leverage the released synthetic data to infer the sensitive attributes that remain unknown. Here, we assume the adversary has the prior knowledge of demographic information and the 100 most frequently occurring concepts for inferring the remaining sensitive information. We randomly sample 5,000 records from the real training dataset and another 5,000 from the synthetic dataset. Using Hamming distance, each real record’s unknown attributes are predicted based on its *K*-nearest neighbors in the synthetic data through majority voting. In this experiment, we set *K* = 1 as in Theodorou *et al*.^22^, and report the mean F1 score of the inferred attributes as the measure of attack success.

A membership inference attack happens when an adversary attempts to infer whether a complete real record they possess is included in the dataset used to train the synthetic data generation model. In this experiment, we simulate such an attack dataset by randomly sampling 5,000 records each from the real training and real test data. For each record in this attack set, we calculate its Hamming distance to its nearest neighbor record in the synthetic dataset. The median of these nearest neighbor distances is used as the cutoff threshold for inference, i.e., any real record with its nearest neighbor distance to the synthetic dataset below the cutoff threshold is predicted to be a member of the real training data. We also experiment with a wide range of distance cutoffs and construct ROC curves in Supplementary Fig. 4.

Nearest-neighbor adversarial accuracy (NNAA) evaluates potential overfitting in the synthetic data generation model by examining the degree to which the generated synthetic data are systematically closer to the real training data than to the real test data. In this experiment, 5,000 records are randomly sampled each from real training (*T* ), real test (*E*) and synthetic dataset (*S*). For each dataset, we calculate the Hamming distance of each record to its nearest neighbor within the same dataset, and the distance to the nearest neighbor in another dataset. We then compare the two distances and calculate the proportion of records that are closer to the same dataset compared to other. For instance, we denote the shortest distance of record *i* in the synthetic dataset to the remaining records as *d*_*SS*_(*i*), and the shortest distance to those in the training dataset as *d*_*ST*_ (*i*), and then calculate the proportion of records in the synthetic dataset that are closer to the same dataset than to the training dataset as:

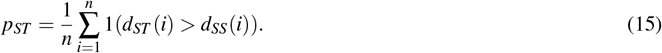

Similarly, we calculate these proportions for *p*_*TS*_, *p*_*SE*_, and *p*_*ES*_. If a model does not overfit to its training data, these quantities are expected to be close to 0.5. Otherwise, *p*_*ST*_ and *p*_*TS*_ would be less than 0.5. Overall, we summarize the risk of overfitting as:

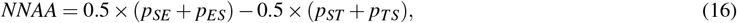

where a positive NNAA value that exceeds a widely accepted threshold (e.g., 0.03 by Yale *et al*.^31^) can raise privacy concerns.

#### Ranking

To summarize each model’s overall strengths and weaknesses across evaluation metrics, we first rank all synthetic datasets generated by the five models (ten replicates per model, totaling 50 synthetic datasets) according to their evaluation results in each metric type. From these data rankings, we then derive normalized model rankings. Lower ranks correspond to higher performance. All model ranks are then linearly normalized to a range from 1 (best) to 5 (worst). Specifically, for each evaluation metric, we rank the 50 synthetic datasets based on their metric values and then compute each model’s rank by averaging the normalized ranks of their ten replicates. For metrics falling within the same type, for example, the outcome prediction performance across three outcomes, we further update each model’s rank as the average across these metrics. Finally, we present model rankings within the utility, fidelity, and privacy dimensions and visualize them using radar plots (Fig. 1e).

## Data Availability

All data produced in the present study are available upon reasonable request to the authors.

## Data Availability

Both the original and synthetic CCASAnet datasets are available upon request. In accordance with the CCASAnet Principles of Collaboration, applicants seeking synthetic data need to submit a concept sheet outlining the intended use of the data. All requests will be reviewed by the CCASAnet Executive Committee. To access the original de-identified CCASAnet data, applicants must complete a more extensive concept sheet and a data use agreement with Vanderbilt University Medical Center. Applications for access to both datasets should be submitted at https://redcap.link/ccasanetsharingrequest. The CCASAnet Executive Committee will start processing applications upon the publication of this study.

## Acknowledgements

This work was supported in part by the NIH-funded Caribbean, Central and South America network for HIV epidemiology (CCASAnet, U01AI069923), by grant R01MH139379 from the National Institute of Mental Health, by grant K99LM014428 from the National Library of Medicine, and by Development Core Award from the Tennessee Center for AIDS Research (TN-CFAR, P30AI110527). The content is solely the responsibility of the authors and does not necessarily represent the official views of the National Institutes of Health. This work is subject to the NIH Public Access Policy. Through acceptance of the federal funding, the NIH has been given the right to make this manuscript publicly available in PubMed Central upon the Official Date of Publication, as defined by the NIH.

## Author Contribution Statement

Z.J.L., C.Y., B.E.S., and B.A.M. conceived the study and designed the experiments. Z.J.L. conducted data preprocessing, performed the experiments, and analyzed the results. Z.L. and N.J. contributed significantly to the model and study design.

Z.J.L. and C.Y. summarized the major experimental findings and drafted the manuscript. B.E.S. and B.A.M. assisted in interpreting the results and extensively revised the manuscript. Z.L., N.J., A.A., A.B., J.M.P., S.N.D., Y.C., R.I., F.P., J.B., D.V., and C.C. provided intellectual input and contributed to manuscript revisions. C.Y., B.E.S., and B.A.M. jointly supervised the study and serve as corresponding authors. All authors participated in manuscript preparation and approved the final version.

