## Supplementary Information for "Generating Synthetic Multi-national Longitudinal Cohorts for Clinically Grounded HIV Research"

| | Argentina (N=1,677) | Brazil (N=10,135) | Chile (N=7,356) | Haiti (N=26,412) | Honduras (N=1,687) | Mexico (N=2,339) | Total (N=49,606) |
|---|---|---|---|---|---|---|---|
| **Year of enrollment** | | | | | | | |
| Mean (SD) | 2007.3 (6.5) | 2010.5 (10.0) | 2013.5 (7.0) | 2012.0 (5.9) | 2009.5 (6.3) | 2011.9 (5.5) | 2011.7 (7.2) |
| Median [IQR] | 2006.0 [2001.0, 2013.0] | 2012.0 [2005.0, 2019.0] | 2015.0 [2009.0, 2019.0] | 2013.0 [2008.0, 2016.0] | 2008.0 [2004.0, 2014.0] | 2013.0 [2008.0, 2016.0] | 2013.0 [2007.0, 2017.0] |
| **Gender** | | | | | | | |
| Female | 421 (25.1%) | 2,668 (26.3%) | 867 (11.8%) | 15,391 (58.3%) | 656 (38.9%) | 252 (10.8%) | 20,255 (40.8%) |
| Male | 1,256 (74.9%) | 7,467 (73.7%) | 6,489 (88.2%) | 11,021 (41.7%) | 1,031 (61.1%) | 2,087 (89.2%) | 29,351 (59.2%) |
| **Age at enrollment** | | | | | | | |
| Mean (SD) | 39.4 (10.5) | 36.1 (11.0) | 34.5 (9.9) | 37.0 (11.1) | 36.2 (10.1) | 36.1 (10.8) | 36.5 (10.9) |
| Median [IQR] | 38.0 [32.0, 46.0] | 34.0 [28.0, 43.0] | 32.0 [27.0, 40.0] | 36.0 [29.0, 44.0] | 35.0 [28.5, 43.0] | 34.0 [28.0, 42.0] | 35.0 [28.0, 43.0] |
| **NO. of clinical endpoints** | | | | | | | |
| Mean (SD) | 2.9 (2.8) | 2.5 (2.4) | 2.3 (2.6) | 1.0 (0.2) | 3.5 (3.4) | 2.5 (2.4) | 1.7 (1.9) |
| Median [IQR] | 2.0 [1.0, 4.0] | 1.0 [1.0, 3.0] | 1.0 [1.0, 3.0] | 1.0 [1.0, 1.0] | 2.0 [1.0, 5.0] | 1.0 [1.0, 3.0] | 1.0 [1.0, 1.0] |
| **NO. of CD4 cell count records** | | | | | | | |
| Mean (SD) | 23.0 (18.3) | 14.7 (16.3) | 7.3 (7.8) | 4.0 (4.9) | 9.5 (8.9) | 17.3 (12.8) | 8.1 (11.3) |
| Median [IQR] | 19.0 [8.0, 37.0] | 8.0 [2.0, 23.0] | 4.0 [2.0, 9.0] | 2.0 [1.0, 5.0] | 6.0 [2.0, 16.0] | 15.0 [6.0, 27.0] | 3.0 [1.0, 10.0] |
| **NO. of HIV viral load records** | | | | | | | |
| Mean (SD) | 23.0 (18.1) | 16.3 (17.4) | 7.1 (7.8) | 4.0 (3.5) | 7.3 (6.8) | 17.5 (12.9) | 8.4 (11.5) |
| Median [IQR] | 18.0 [8.0, 37.0] | 9.0 [2.0, 26.0] | 4.0 [2.0, 9.0] | 2.0 [1.0, 7.0] | 5.0 [1.0, 12.0] | 15.0 [7.0, 26.5] | 4.0 [1.0, 9.0] |
| **Total follow-up (year)** | | | | | | | |
| Mean (SD) | 9.5 (7.2) | 7.6 (7.5) | 4.4 (5.2) | 6.8 (6.1) | 7.8 (6.7) | 7.7 (5.7) | 6.8 (6.4) |
| Median [IQR] | 8.0 [3.0, 15.0] | 5.0 [1.0, 13.0] | 3.0 [1.0, 6.0] | 5.0 [2.0, 11.0] | 6.0 [1.5, 14.0] | 7.0 [3.0, 12.0] | 5.0 [1.0, 11.0] |
| **Death** | | | | | | | |
| Alive/Censored | 1,611 (96.1%) | 7,841 (77.4%) | 6,757 (91.9%) | 23,444 (88.8%) | 1,290 (76.5%) | 2,040 (87.2%) | 42,983 (86.6%) |
| Death | 66 (3.9%) | 2,294 (22.6%) | 599 (8.1%) | 2,968 (11.2%) | 397 (23.5%) | 299 (12.8%) | 6,623 (13.4%) |

**Supplementary Table 1.** Summary statistics of the CCASAnet cohort of PWH used in this study.

| Table Name | Variable | Content |
| --- | --- | --- |
| Basic | Person ID | Unique person identifier, e.g. ‘123’. |
| | Gender | Gender at enrollment, either ‘male’ or ‘female’. |
| | Site | Country of HIV clinic for person enrollment, e.g. ‘Haiti’. |
| | Transmission mode | Mode of HIV infection, e.g., ‘MSM’. |
| | Date of birth | Date of birth, e.g., ‘1990-01-01’. |
| | Date of enrollment | Date of enrollment in the HIV clinic. |
| | Date of HIV diagnosis | Date of HIV virus detection. |
| Visit | Person ID | Unique person identifier. |
| | Date of visit | Date of clinical visit. |
| | Weight | Weight measured during visit, e.g., 100kg. |
| | Height | Height measured during visit, e.g., 160cm. |
| | WHO HIV stage classification | Current stage in WHO HIV and AIDS classification. |
| | CDC HIV stage classification | Current stage in CDC HIV and AIDS classification. |
| Clinical endpoint | Person ID | Unique person identifier. |
| | Date of diagnosis | Date of clinical endpoint diagnosis. |
| | Clinical endpoint | Name of clinical endpoint; if it is an AIDS defining event. |
| CD4 | Person ID | Unique person identifier. |
| | Date of CD4 cell measurement | Date of CD4 cell count measurement. |
| | CD4 cell count | The number of CD4 cell measured, e.g., 300. |
| HIV viral load | Person ID | Unique person identifier. |
| | Date of HIV viral load measurement | Date of the HIV viral load measurement. |
| | HIV viral load | Measurement of HIV viral load, e.g., 100,000. |
| Medication | Person ID | Unique person identifier. |
| | Starting date of ART treatment | Starting date of current treatment. |
| | Ending date of ART treatment | Ending date of current treatment, can be empty. |
| | ART regimen | Name of ART regimens combination, e.g., ‘3TC,AZT,EFV’. |
| | Highly active ART classification | If this ART combination is highly active. |
| | RTV drug | If the regimen contains any RTV drug. |
| Follow-up | Person ID | Unique person identifier. |
| | Last observation date | Latest date on which the person was observed. |
| | Dead | If the person died. |
| | Lost-to-follow-up | If the person was lost to follow-up. |

**Supplementary Table 2.** A summary of CCASAnet variables and their source tables used in this study.

| **Model** | **Log-rank test $p$-value** | **KM-D** | **Maximum follow-up time (years)** |
|---|---|---|---|
| Real training (vs. real test) | 0.750 | 0.035 | 35.965 (vs. 35.663) |
| MeLD | **0.815 [0.521, 1.000]** | **0.015 [0.005, 0.024]** | **38.884 [35.451, 42.317]** |
| SynTEG | 0.192 [0.000, 0.576] | 0.048 [0.040, 0.056] | 92.102 [76.578, 107.627] |
| HALO | 0.000 [0.000, 0.000] | 0.055 [0.016, 0.094] | 45.451 [37.922, 52.981] |
| TimeDiff | 0.000 [0.000, 0.000] | 0.080 [0.056, 0.104] | 93.342 [59.762, 126.921] |
| TimeAutoDiff | 0.199 [0.000, 0.680] | 0.055 [0.000, 0.135] | 42.785 [37.177, 48.392] |
| MeLD-DDPM | 0.000 [0.000, 0.000] | 0.106 [0.095, 0.117] | 42.563 [35.477, 49.650] |
| MeLD-LSTM | 0.034 [0.000, 0.111] | 0.045 [0.038, 0.052] | 53.681 [45.494, 61.868] |
| MeLD-DiT | 0.000 [0.000, 0.000] | 0.128 [0.120, 0.137] | 251.530 [191.468, 311.591] |
| MeLD-Transformer | **0.417 [0.000, 0.855]** | **0.018 [0.009, 0.028]** | **37.958 [34.803, 41.112]** |

**Supplementary Table 3.** Performance comparison in time-to-death estimations. Log-rank test $p$-values, Kaplan-Meier distance (KM-D), and maximum follow-up for time from ART initiation to death are derived using real test data as the reference. Results based on real training data serve as the baseline. 95% confidence intervals are reported over ten synthetically generated datasets. Best-performing results are highlighted in bold for both comparisons between MeLD and existing models and comparisons among MeLD variants.

| | **Prediction task** | | |
|---|---|---|---|
| **Model** | **Tuberculosis** | **Cancer** | **Cardiovascular disease** |
| Real | 0.763 [0.752, 0.774] | 0.616 [0.596, 0.636] | 0.659 [0.642, 0.676] |
| MeLD | **0.757 [0.741, 0.773]** | **0.626 [0.597, 0.655]** | <u>0.615 [0.541, 0.689]</u> |
| SynTEG | <u>0.756 [0.745, 0.767]</u> | 0.595 [0.545, 0.645] | 0.598 [0.534, 0.662] |
| HALO | 0.745 [0.728, 0.762] | <u>0.618 [0.579, 0.657]</u> | 0.559 [0.469, 0.649] |
| TimeDiff | 0.752 [0.736, 0.768] | 0.599 [0.571, 0.627] | **0.639 [0.566, 0.712]** |
| TimeAutoDiff | 0.748 [0.737, 0.759] | 0.603 [0.584, 0.622] | 0.531 [0.464, 0.598] |
| MeLD-DDPM | 0.719 [0.707, 0.731] | 0.591[0.577, 0.605] | 0.577 [0.573, 0.582] |
| MeLD-LSTM | 0.747 [0.741, 0.754] | 0.598 [0.558, 0.637] | 0.571 [0.487, 0.654] |
| MeLD-DiT | 0.496 [0.374, 0.619] | 0.544 [0.476, 0.613] | 0.540 [0.392, 0.693] |
| MeLD-Transformer | **0.761 [0.745, 0.776]** | **0.627 [0.590, 0.663]** | **0.642 [0.603, 0.680]** |

**Supplementary Table 4.** Prediction performance on the real test dataset characterized by AUROC for tuberculosis, cancer, and cardiovascular diseases. Models are trained on information accumulated during the first six months after enrollment to predict subsequent clinical endpoints. The scenario of TRTR (i.e., Real) serves as the reference. 95% confidence intervals are reported over ten synthetically generated datasets, whereas those of TRTR are based on random splits of training and validation sets. Best-performing results are highlighted in bold for both comparisons between MeLD and existing models and comparisons among MeLD variants. Second-best results are underlined in comparisons between MeLD and existing models.

| Risk factor | Model | Pooled estimate | Bias | SE ratio | CI overlap | Type I/ II err. | Direction err. |
|---|---|---|---|---|---|---|---|
| Age at ART initiation (30 vs. 20) | Real Training | 0.069 [-0.001, 0.139] | - | - | - | - | - |
| | MeLD | 0.035 [-0.045,0.116] | -0.034 | 1.157 | 0.565 | 1 / - | - |
| | SynTEG | 0.147 [0.076, 0.218] | 0.078 | 1.025 | 0.292 | 9 / - | - |
| | HALO | 0.024 [-0.035, 0.083] | -0.045 | 0.848 | 0.490 | 1 / - | - |
| | TimeDiff | 0.038 [-0.071, 0.147] | -0.031 | 1.561 | 0.511 | 1 / - | - |
| | TimeAutoDiff | 0.072 [-0.007,0.152] | 0.003 | 1.143 | 0.526 | 5 / - | - |
| | MeLD-DDPM | 0.258 [0.134,0.382] | 0.189 | 1.779 | 0.096 | 10 / - | - |
| | MeLD-LSTM | -0.037 [-0.112,0.038] | -0.106 | 1.079 | 0.191 | 2 / - | - |
| | MeLD-DiT | -0.003 [-0.150,0.144] | -0.072 | 2.111 | 0.387 | 1 / - | - |
| | MeLD-Transformer | -0.007 [-0.086,0.073] | -0.076 | 1.142 | 0.338 | 1 / - | - |
| CD4 cell count at ART initiation (200 vs. 100) | Real training | -0.223 [-0.320,-0.126]* | - | - | - | - | - |
| | MeLD | -0.422 [-0.553,-0.290] | -0.199 | 1.360 | 0.131 | - / 0 | 0 |
| | SynTEG | -0.332 [-0.422,-0.242] | -0.110 | 0.929 | 0.283 | - / 0 | 0 |
| | HALO | -0.053 [-0.104,-0.002] | 0.170 | 0.530 | 0.015 | - / 4 | 0 |
| | TimeDiff | -0.083 [-0.157,-0.009] | 0.140 | 0.763 | 0.141 | - / 4 | 0 |
| | TimeAutoDiff | 0.054 [-0.050,0.158] | 0.277 | 1.075 | 0.000 | - / 9 | 1 |
| | MeLD-DDPM | -0.279 [-0.410,-0.148] | -0.056 | 1.347 | 0.507 | - / 0 | 0 |
| | MeLD-LSTM | -0.285 [-0.421,-0.149] | -0.063 | 1.404 | 0.519 | - / 0 | 0 |
| | MeLD-DiT | 0.035 [0.024,0.047] | 0.258 | 0.121 | 0.000 | - / 0 | 10 |
| | MeLD-Transformer | -0.445 [-0.588,-0.303] | -0.223 | 1.471 | 0.067 | - / 0 | 0 |
| Calendar year of ART initiation (2020 vs. 2010) | Real training | 0.310 [0.226, 0.394]* | - | - | - | - | - |
| | MeLD | 1.250 [1.009,1.490] | 0.939 | 2.860 | 0.000 | - / 0 | 0 |
| | SynTEG | -0.418 [-0.513, -0.323] | -0.729 | 1.126 | 0.000 | - / 0 | 0 |
| | HALO | -0.139 [-0.266, -0.012] | -0.449 | 1.515 | 0.000 | - / 4 | 6 |
| | TimeDiff | 0.156 [-0.274, 0.586] | -0.155 | 5.123 | 0.182 | - / 9 | 0 |
| | TimeAutoDiff | 0.046 [-0.064,0.156] | -0.264 | 1.314 | 0.029 | - / 6 | 0 |
| | MeLD-DDPM | 1.095 [0.794,1.397] | 0.785 | 3.592 | 0.000 | - / 0 | 0 |
| | MeLD-LSTM | -0.033 [-0.216,0.150] | -0.343 | 2.176 | 0.025 | - / 9 | 1 |
| | MeLD-DiT | -0.001 [-0.149,0.146] | -0.312 | 1.753 | 0.012 | - / 10 | 0 |
| | MeLD-Transformer | -0.201 [-0.432,0.030] | -0.511 | 2.750 | 0.000 | - / 7 | 3 |
| Gender (Male vs. Female) | Real training | 0.068 [0.010, 0.126]* | - | - | - | - | - |
| | MeLD | 0.120 [0.057,0.183] | 0.052 | 1.079 | 0.463 | - / 0 | 0 |
| | SynTEG | 0.076 [0.013, 0.139] | 0.008 | 1.071 | 0.723 | - / 4 | 0 |
| | HALO | 0.193 [0.133, 0.253] | 0.125 | 1.035 | 0.044 | - / 0 | 0 |
| | TimeDiff | 0.079 [0.008, 0.150] | 0.011 | 1.213 | 0.642 | - / 4 | 0 |
| | TimeAutoDiff | -0.134 [-0.211,-0.057] | -0.202 | 1.322 | 0.022 | - / 1 | 9 |
| | MeLD-DDPM | 0.238 [0.155,0.321] | 0.170 | 1.417 | 0.036 | - / 0 | 0 |
| | MeLD-LSTM | 0.021 [-0.044,0.086] | -0.047 | 1.106 | 0.421 | - / 9 | 0 |
| | MeLD-DiT | 0.075 [-0.092,0.241] | 0.007 | 2.854 | 0.320 | - / 8 | 0 |
| | MeLD-Transformer | 0.117 [0.055,0.179] | 0.049 | 1.068 | 0.467 | - / 0 | 0 |

**Supplementary Table 5.** Comparison of log-hazard ratios for mortality risk factors at ART initiation between synthetic data and real data. For each risk factor, Cox-proportional hazard estimates from real training data are shown in the first row as the reference, with significant estimates marked with asterisks. The summary columns from left to right are: 1) pooled estimate: combined estimate of raw estimates from synthetic data replicates using Rubin's rule; 2) bias: mean difference of estimates obtained from synthetic data and that from real data; 3) SE ratio: mean estimates' standard errors (SE) from synthetic data over that from real data; 4) CI overlap: mean proportions of confidence interval (CI) from real data covered by CIs from synthetic data, normalized by the width of union of both CIs; 5) type I/II err: counts of type I or II errors; 6) direction err: counts of direction errors.

| Model | Visit number | Time gap | CD4 cell count | HIV viral load | Enrollment age | Height | Weight |
|---|---|---|---|---|---|---|---|
| MeLD | **0.507**<br>[0.330, 0.685] | **0.073**<br>[0.070, 0.075] | 1.173<br>[1.139, 1.207] | **0.138**<br>[0.123, 0.152] | 0.535<br>[0.505, 0.566] | **0.062**<br>[0.061, 0.063] | **1.939**<br>[1.852, 2.026] |
| SynTEG | 22.199<br>[21.926, 22.473] | 0.665<br>[0.643, 0.687] | 2.964<br>[2.928, 3.001] | 0.656<br>[0.652, 0.660] | **0.177**<br>[0.173, 0.180] | 0.079<br>[0.078, 0.079] | 4.184<br>[4.043, 4.325] |
| HALO | 1.553<br>[1.129, 1.977] | 0.128<br>[0.119, 0.136] | 2.974<br>[2.886, 3.061] | 0.485<br>[0.455, 0.515] | 8.000<br>[7.857, 8.142] | 0.094<br>[0.094, 0.095] | 3.787<br>[3.737, 3.836] |
| TimeDiff | 20.454<br>[20.128, 20.780] | 0.214<br>[0.199, 0.229] | 3.774<br>[3.513, 4.034] | 1.048<br>[1.015, 1.082] | 16.966<br>[16.853, 17.080] | 0.172<br>[0.170, 0.174] | 6.231<br>[6.197, 6.265] |
| TimeAutoDiff | 5.222<br>[4.870, 5.574] | 0.215<br>[0.196, 0.234] | **0.959**<br>[0.900, 1.019] | 0.437<br>[0.403, 0.471] | 17.812<br>[17.759, 17.865] | 0.102<br>[0.079, 0.125] | 2.015<br>[1.859, 2.171] |
| MeLD-DDPM | **0.543**<br>[0.481, 0.605] | 0.130<br>[0.128, 0.132] | 1.142<br>[1.107, 1.177] | 0.851<br>[0.822, 0.880] | 1.754<br>[1.713, 1.795] | **0.037**<br>[0.036, 0.038] | 5.351<br>[5.318, 5.385] |
| MeLD-LSTM | 8.056<br>[7.870, 8.241] | 0.131<br>[0.129, 0.133] | 0.805<br>[0.775, 0.835] | 0.398<br>[0.379, 0.416] | 1.118<br>[1.077, 1.159] | 0.059<br>[0.058, 0.061] | 2.569<br>[2.530, 2.608] |
| MeLD-DiT | 32.578<br>[32.505, 32.651] | 1.961<br>[1.953, 1.969] | 13.165<br>[12.928, 13.401] | 6.393<br>[6.356, 6.431] | 11.728<br>[11.538, 11.917] | 0.621<br>[0.607, 0.636] | 27.115<br>[26.140, 28.090] |
| MeLD-Transformer | 1.006<br>[0.816, 1.196] | **0.087**<br>[0.084, 0.089] | **0.656**<br>[0.623, 0.689] | **0.374**<br>[0.344, 0.403] | **0.781**<br>[0.742, 0.820] | 0.065<br>[0.061, 0.069] | **0.551**<br>[0.492, 0.609] |

**Supplementary Table 6.** Fidelity comparison in continuous variables between synthetic and real data. Fidelity is characterized by Wasserstein distance (WSD) between the variable distribution obtained from synthetic data and that from real data. Mean and 95% confidence intervals of each continuous variable are reported over ten synthetic datasets. Best-performing results are highlighted in bold for both comparisons between MeLD and existing models and comparisons among MeLD variants. The second best results are underlined for comparison between MeLD and existing models.

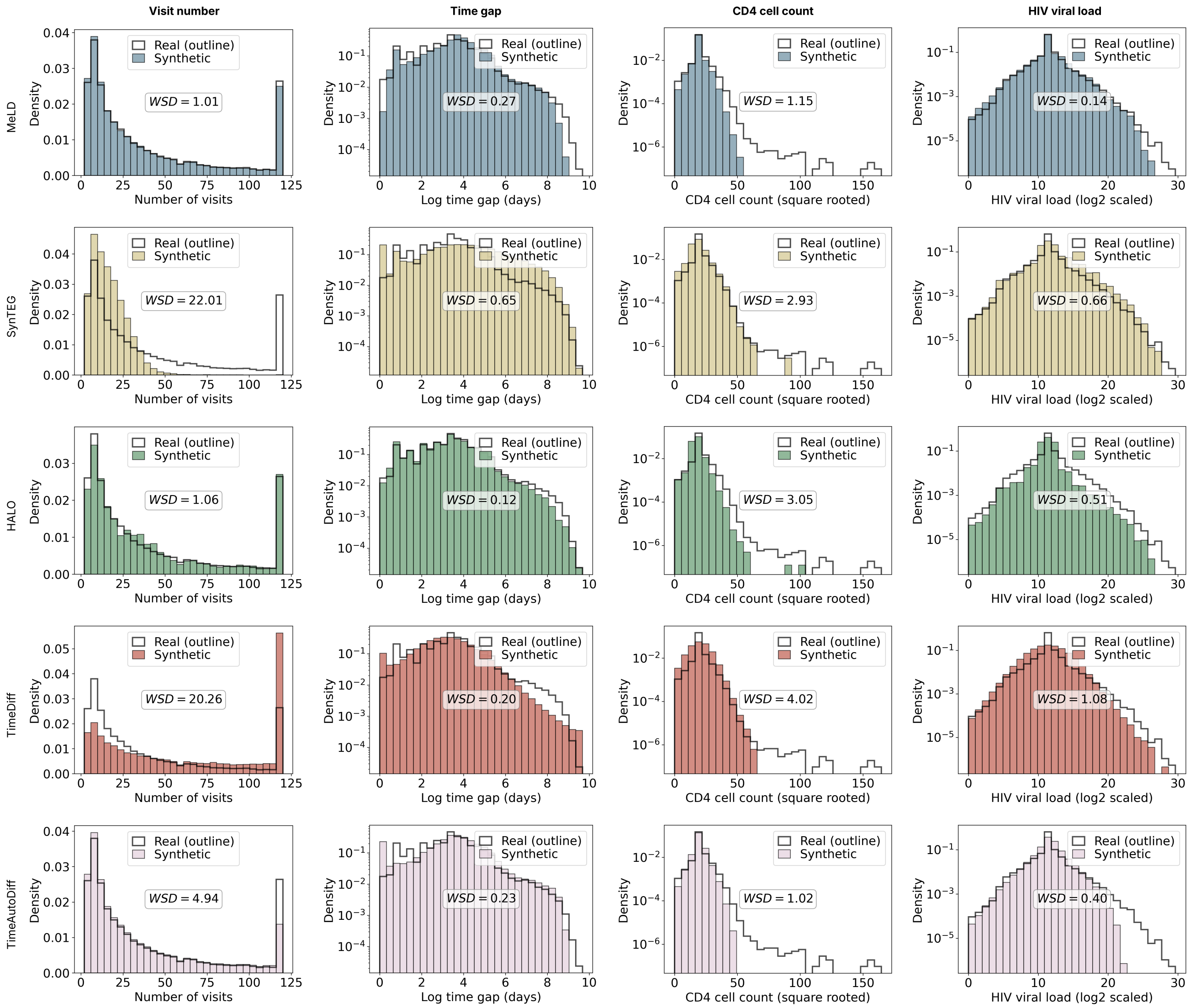


**Supplementary Figure 1.** Distribution comparison between the distributions of continuous variables derived from real data and a representative synthetic dataset, selected to be closest to the mean Wasserstein distance (WSD) across all replicates. Distributions obtained from real data are displayed as black outlines, while those from synthetic data are shown as solid bars.

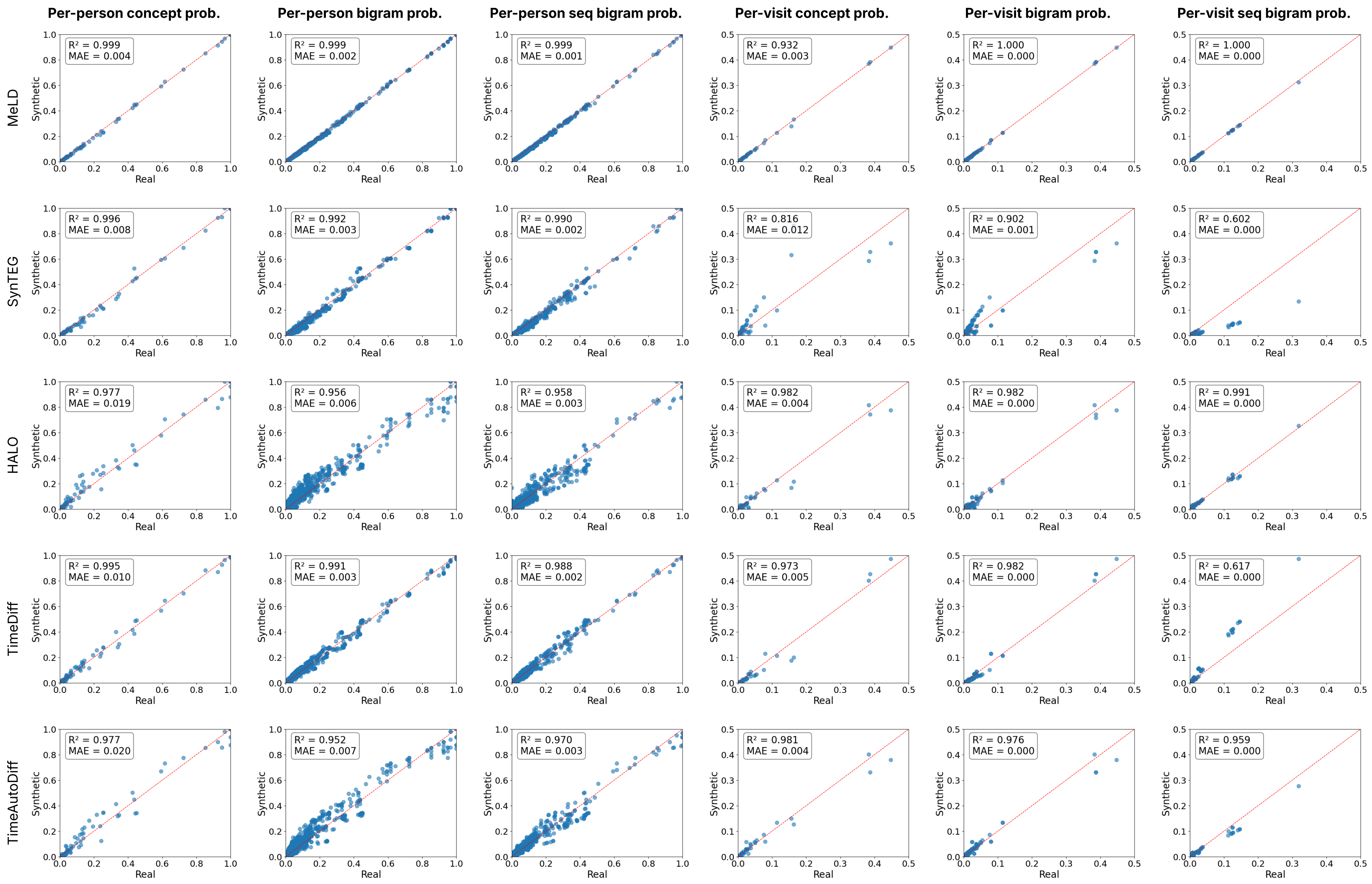


**Supplementary Figure 2.** Fidelity comparison between the distributions of categorical variables obtained from real data and those from a representative synthetic dataset, selected to be closest to the mean absolute error (MAE) across all replicates. Categorical variables or their pairs are shown as scatterplot, with x- and y-axes denoting prevalence in real and synthetic data respectively. Both MAE and $R^2$ values are indicated.

| Categorical variable distribution metric | Model | Mean absolute error | $R^2$ |
|---|---|---|---|
| Per-person concept probability | SynTEG | 0.0074 [0.0067, 0.0081] | 0.996 [0.996, 0.997] |
| | HALO | 0.0192 [0.0179, 0.0205] | 0.976 [0.973, 0.980] |
| | TimeDiff | 0.0106 [0.0103, 0.0110] | 0.995 [0.994, 0.995] |
| | TimeAutoDiff | 0.0172 [0.0120, 0.0224] | 0.982 [0.973, 0.991] |
| | MeLD-DDPM | 0.0184 [0.0181, 0.0187] | 0.983 [0.982, 0.983] |
| | MeLD-LSTM | 0.0215 [0.0213, 0.0218] | 0.964 [0.963, 0.965] |
| | MeLD-DiT | 0.1264 [0.1259, 0.1269] | 0.463 [0.460, 0.467] |
| | MeLD-Transformer | 0.0048 [0.0045, 0.0051] | 0.999 [0.999, 0.999] |
| | **MeLD** | **0.0038 [0.0035, 0.0040]** | 0.999 [0.999, 0.999] |
| Per-person bigram probability | SynTEG | 0.0030 [0.0029, 0.0031] | 0.992 [0.991, 0.993] |
| | HALO | 0.0059 [0.0053, 0.0066] | 0.954 [0.945, 0.962] |
| | TimeDiff | 0.0035 [0.0034, 0.0036] | 0.990 [0.990, 0.991] |
| | TimeAutoDiff | 0.0061 [0.0040, 0.0083] | 0.964 [0.941, 0.987] |
| | MeLD-DDPM | 0.0071 [0.0070, 0.0072] | 0.965 [0.964, 0.966] |
| | MeLD-LSTM | 0.0080 [0.0078, 0.0082] | 0.925 [0.924, 0.927] |
| | MeLD-DiT | 0.0442 [0.0432, 0.0453] | -0.008 [-0.036, 0.021] |
| | MeLD-Transformer | 0.0020 [0.0019, 0.0022] | 0.998 [0.998, 0.998] |
| | **MeLD** | **0.0016 [0.0015, 0.0017]** | 0.999 [0.998, 0.999] |
| Per-person sequential bigram probability | SynTEG | 0.0015 [0.0013, 0.0017] | 0.991 [0.988, 0.993] |
| | HALO | 0.0030 [0.0028, 0.0033] | 0.955 [0.946, 0.964] |
| | TimeDiff | 0.0021 [0.0020, 0.0021] | 0.987 [0.986, 0.988] |
| | TimeAutoDiff | 0.0026 [0.0024, 0.0029] | 0.976 [0.965, 0.987] |
| | MeLD-DDPM | 0.0037 [0.0037, 0.0038] | 0.963 [0.962, 0.964] |
| | MeLD-LSTM | 0.0043 [0.0042, 0.0043] | 0.907 [0.904, 0.909] |
| | MeLD-DiT | 0.0119 [0.0117, 0.0122] | 0.314 [0.301, 0.327] |
| | MeLD-Transformer | 0.0011 [0.0010, 0.0012] | 0.997 [0.997, 0.998] |
| | **MeLD** | **0.0008 [0.0008, 0.0009]** | 0.999 [0.998, 0.999] |
| Per-visit concept probability | SynTEG | 0.0128 [0.0107, 0.0149] | 0.804 [0.754, 0.853] |
| | HALO | 0.0038 [0.0032, 0.0044] | 0.980 [0.971, 0.990] |
| | TimeDiff | 0.0046 [0.0042, 0.0050] | 0.977 [0.971, 0.983] |
| | TimeAutoDiff | 0.0036 [0.0031, 0.0040] | 0.984 [0.978, 0.989] |
| | MeLD-DDPM | 0.0067 [0.0064, 0.0071] | 0.900 [0.882, 0.918] |
| | MeLD-LSTM | 0.0119 [0.0116, 0.0122] | 0.706 [0.688, 0.724] |
| | MeLD-DiT | 0.0434 [0.0432, 0.0436] | 0.137 [0.127, 0.146] |
| | MeLD-Transformer | 0.0027 [0.0024, 0.0031] | 0.954 [0.938, 0.970] |
| | **MeLD** | **0.0025 [0.0021, 0.0029]** | 0.951 [0.928, 0.974] |
| Per-visit bigram probability | SynTEG | 0.0008 [0.0006, 0.0010] | 0.879 [0.830, 0.928] |
| | HALO | 0.0003 [0.0003, 0.0003] | 0.973 [0.950, 0.995] |
| | TimeDiff | 0.0003 [0.0003, 0.0003] | 0.981 [0.978, 0.983] |
| | TimeAutoDiff | 0.0002 [0.0002, 0.0003] | 0.979 [0.973, 0.985] |
| | MeLD-DDPM | 0.0004 [0.0004, 0.0004] | 0.908 [0.900, 0.915] |
| | MeLD-LSTM | 0.0007 [0.0007, 0.0007] | 0.583 [0.573, 0.593] |
| | MeLD-DiT | 0.0037 [0.0036, 0.0037] | -1.000 [-1.021, -0.979] |
| | MeLD-Transformer | **0.0001 [0.0001, 0.0001]** | 0.998 [0.997, 1.000] |
| | **MeLD** | **0.0001 [0.0001, 0.0001]** | 1.000 [0.999, 1.000] |
| Per-visit sequential bigram probability | SynTEG | 0.0002 [0.0002, 0.0002] | 0.585 [0.550, 0.621] |
| | HALO | 0.0001 [0.0001, 0.0001] | 0.984 [0.969, 0.999] |
| | TimeDiff | 0.0002 [0.0002, 0.0002] | 0.608 [0.588, 0.628] |
| | TimeAutoDiff | 0.0001 [0.0001, 0.0001] | 0.966 [0.953, 0.978] |
| | MeLD-DDPM | 0.0001 [0.0001, 0.0001] | 0.938 [0.933, 0.943] |
| | MeLD-LSTM | 0.0003 [0.0003, 0.0003] | 0.596 [0.586, 0.606] |
| | MeLD-DiT | 0.0003 [0.0003, 0.0003] | 0.173 [0.170, 0.177] |
| | MeLD-Transformer | **0.0000 [0.0000, 0.0000]** | 0.999 [0.998, 0.999] |
| | **MeLD** | **0.0000 [0.0000, 0.0000]** | 1.000 [0.999, 1.000] |

**Supplementary Table 7.** Fidelity comparison between the distributions of categorical variables obtained from real data and those from synthetic data. 95% confidence intervals are reported over ten synthetically generated datasets. Best-performing MAE results are highlighted in bold.

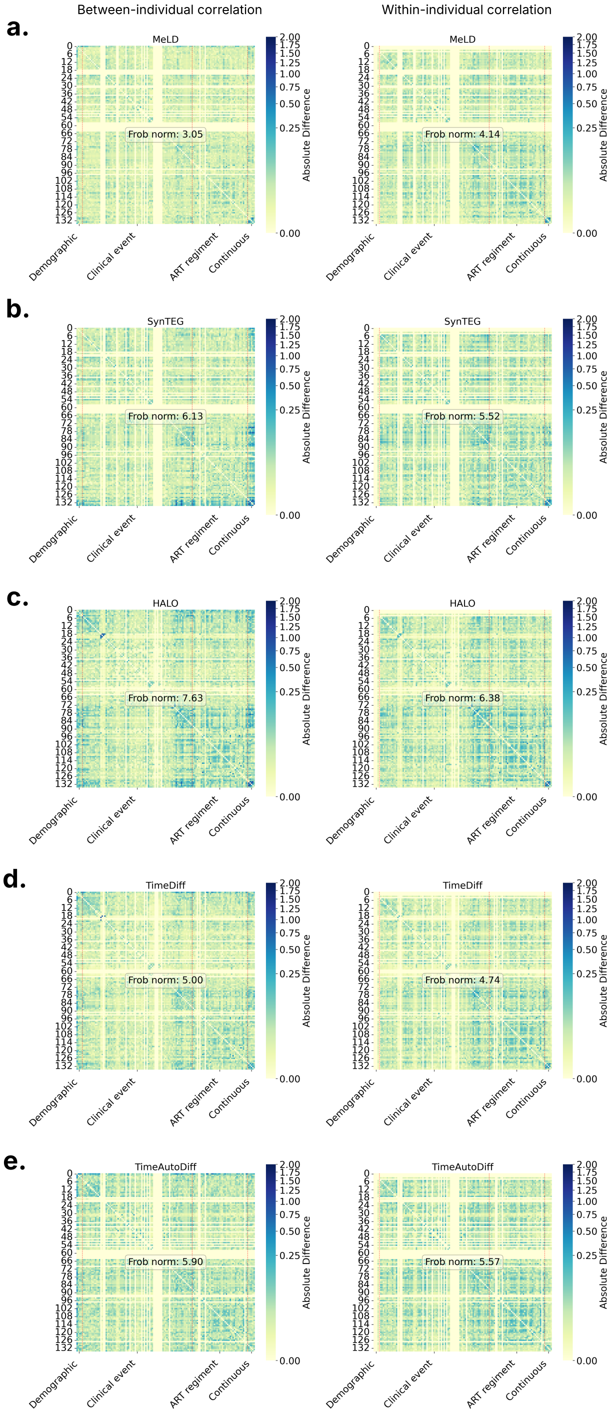


**Supplementary Figure 3.** Differences between variable correlations derived from real data and those from a representative synthetic dataset, selected to be closest to the mean Frobenius norm across all replicates. Variables' correlations, including between- and within-individual correlations, are calculated using Spearman's rank correlation. The total difference is characterized with Frobenius norm. The entries of each heatmap are grouped as demographics, clinical events, antiretroviral therapy (ART) regimen, and continuous variables, which are separated using dotted red lines.

| Model | Normalized mutual information |
|---|---|
| Test (vs. real training) | 0.0001 [0.0000,0.0005] |
| MeLD | **0.0003 [0.0000,0.0009]** |
| SynTEG | 0.1117 [0.0399,0.1834] |
| HALO | 0.0012 [0.0000,0.0040] |
| TimeDiff | 0.0377 [0.0317,0.0437] |
| TimeAutoDiff | 0.0013 [0.0000,0.0029] |
| MeLD-DDPM | 0.0021 [0.0000,0.0062] |
| MeLD-LSTM | 0.0218 [0.0053,0.0384] |
| MeLD-DiT | 0.0988 [0.0000,0.2218] |
| MeLD-Transformer | **0.0004 [0.0000,0.0010]** |

**Supplementary Table 8.** Fidelity comparison between latent clustering consistency derived from synthetic data and that from real data. Latent clustering consistency is characterized by normalized mutual information (NMI) between data labels (e.g., real vs. synthetic) and cluster labels. Analysis with NMI close to 0 indicates inseparability between synthetic and real data in the latent space. Results based on “test vs. real training” serves as the reference. 95% confidence intervals are reported over ten synthetically generated datasets, whereas the one for “test vs. real training” is based on random samples of real data. Best-performing results are highlighted in bold for both comparisons between MeLD and alternative models and comparisons among MeLD variants.

| Model | Attribution inference | Membership inference | NNAA |
|---|---|---|---|
| | F1 score | Accuracy | - |
| MeLD | 0.031 [0.026, 0.035] | **0.509 [0.507, 0.511]** | **-0.0008 [-0.0024, 0.0008]** |
| SynTEG | 0.042 [0.031, 0.052] | 0.510 [0.507, 0.512] | -0.0003 [-0.0018, 0.0013] |
| HALO | 0.049 [0.041, 0.058] | 0.519 [0.506, 0.531] | 0.0188 [-0.0512, 0.0888] |
| TimeDiff | **0.028 [0.025, 0.030]** | 0.511 [0.510, 0.513] | 0.0309 [0.0123, 0.0495] |
| TimeAutoDiff | 0.031 [0.030, 0.033] | **0.509 [0.509, 0.510]** | -0.0001 [-0.0005, 0.0003] |
| MeLD-DDPM | 0.024 [0.021, 0.027] | 0.510 [0.508, 0.511] | -0.0005 [-0.0009, -0.0001] |
| MeLD-LSTM | **0.022 [0.015, 0.028]** | 0.510 [0.508, 0.511] | -0.0002 [-0.0011, 0.0008] |
| MeLD-DiT | 0.037 [0.032, 0.042] | 0.510 [0.509, 0.511] | **-0.0037 [-0.0049, -0.0026]** |
| MeLD-Transformer | 0.024 [0.021, 0.027] | 0.510 [0.509, 0.511] | -0.0001 [-0.0005, 0.0004] |

**Supplementary Table 9.** Privacy risks for attribution inference, membership inference, and nearest neighbor adversarial accuracy (NNAA). 95% confidence intervals are reported over ten synthetically generated datasets. Best-performing results are highlighted in bold for both comparisons between MeLD and alternative models and comparisons among MeLD variants.

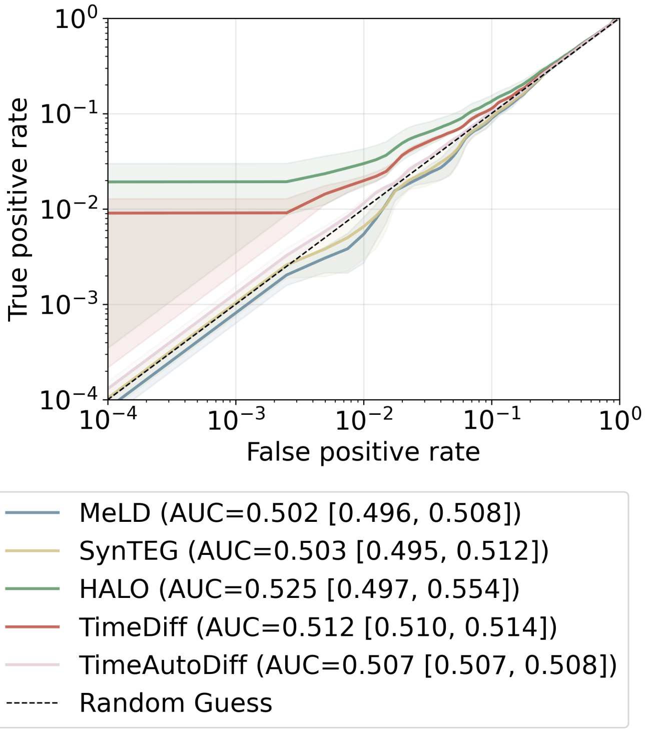


**Supplementary Figure 4.** Sensitivity analysis of membership inference risk. Both x- and y-axes are log-scaled to emphasize membership inference risk in the critical region where the false positive rate is low. 95% confidence intervals are report over ten synthetic generated datasets.
